# A Bibliometric and Visual Analysis of Global Research Trends in the Immunogenomics of *Leishmania spp.* and *Toxoplasma gondii* within a One Health Framework

**DOI:** 10.1101/2025.11.24.25340873

**Authors:** Saqib Shamim, Faraz Shamim

## Abstract

**Background:** Leishmaniasis and toxoplasmosis are major neglected zoonotic diseases affecting billions globally, with diverse clinical outcomes driven by complex host-pathogen immunogenomic interactions. While the One Health framework is theoretically essential for controlling these environmentally sensitive pathogens, the extent of its actual integration into high-throughput genomic research remains unquantified. A systematic mapping of this field is necessary to identify structural gaps and guide future interdisciplinary strategies.

**Objective:** This study aims to conduct a comprehensive bibliometric and visual analysis of global research trends in the immunogenomics of *Leishmania spp.* and *Toxoplasma gondii*, explicitly evaluating the integration of human, animal, and environmental health dimensions within a One Health context.

**Methods:** A systematic analysis was performed on a curated, deduplicated dataset of 1,320 publications retrieved from Dimensions and Lens.org, covering the period from January 2000 to October 2025. Advanced scientometric techniques, including social network analysis, keyword co-occurrence mapping, and automated content classification, were executed using Python to quantify publication trajectories, collaboration networks, and thematic evolution.

**Results:** The research domain exhibited robust exponential growth with a Compound Annual Growth Rate (CAGR) of 8.87% and a doubling time of 5.5 years. Brazil emerged as the leading contributor (142 publications), while the United States led in citation impact. Network analysis revealed a “small-world” collaboration structure (clustering coefficient 0.919) driven by strategic North-South partnerships, particularly between the USA and Brazil. Although 62.9% of publications were classified as integrating One Health concepts, a distinct “environmental blind spot” was identified, with only 5.9% of research addressing the human-environment interface. Thematically, the field has pivoted from observational cytokine profiling (dominated by IL-10 and IFN-γ) to systems-level inquiry, with “molecular docking simulation” (Burst Score 72.7) emerging as a high-velocity frontier, signaling a rapid shift toward *in silico* therapeutic discovery.

**Conclusions:** The “genomic revolution” has successfully democratized research capacity, shifting the center of knowledge production toward endemic regions. However, the field faces a critical technological lag in Artificial Intelligence adoption and a lack of mechanistic environmental research. Future strategies must prioritize “eco-immunogenomics” and multi-omics integration to bridge the gap between genomic data generation and holistic One Health implementation.

**Highlights:**

- Research output grew exponentially (CAGR 8.87%) with a 5.5-year doubling time.
- Brazil leads global production, challenging traditional Global North dominance.
- Strategic North-South partnerships, notably USA-Brazil, drive global collaboration.
- Only ∼6% of publications address the environmental component of One Health.
- In silico docking is a rapid frontier, yet AI adoption remains a critical gap.

## 1. Introduction

### Background: The Persistent Global Challenge of Leishmaniasis and Toxoplasmosis

Leishmaniasis and toxoplasmosis, two protozoan parasitic diseases of profound global significance, collectively impose a staggering burden on human health and socioeconomic development. Leishmaniasis, a vector-borne disease caused by over 20 species of the genus *Leishmania*, is transmitted by the bite of infected phlebotomine sandflies. The World Health Organization reports that the disease is endemic in approximately 90 countries across Asia, Africa, the Americas, and the Mediterranean basin, placing over one billion people at risk. The clinical spectrum of leishmaniasis is remarkably diverse, ranging from self-healing cutaneous ulcers (cutaneous leishmaniasis, CL), which can result in disfiguring scars and significant social stigma, to the life-threatening visceral leishmaniasis (VL), also known as kala-azar (Burza et al., 2018). VL is the most severe form, characterized by prolonged fever, weight loss, hepatosplenomegaly, and pancytopenia. Without treatment, VL is fatal in over 95% of cases, causing tens of thousands of deaths annually, primarily among the world’s most impoverished populations (Burza et al., 2018).

Concurrently, toxoplasmosis, caused by the cosmopolitan protozoan *Toxoplasma gondii*, stands as one of the most successful parasitic infections, with an estimated one-third of the global human population carrying a chronic infection (Pappas et al., 2009). Transmission primarily occurs through the ingestion of food or water contaminated with oocysts shed by felids; the only definitive hosts; or through the consumption of undercooked meat containing parasitic tissue cysts. While typically asymptomatic in immunocompetent individuals, *T. gondii* infection can lead to severe, life-threatening complications, including encephalitis and myocarditis, in immunocompromised patients, such as those with HIV/AIDS or organ transplant recipients (Montoya & Liesenfeld, 2004). The most devastating consequence is congenital toxoplasmosis, which arises from primary maternal infection during pregnancy and can result in miscarriage, stillbirth, or severe neurological and ocular disease in the newborn, contributing to a substantial global burden of disability (Torgerson & Mastroiacovo, 2013).

The effective control of both leishmaniasis and toxoplasmosis is hampered by a formidable array of challenges. Diagnostic tools for both diseases often lack the requisite sensitivity and specificity, or are too complex and costly for implementation in the resource-limited settings where they are most needed (Cunningham et al., 2019). Therapeutic arsenals are limited, frequently associated with significant toxicity, and are increasingly threatened by the emergence of drug resistance. The rise of resistance to pentavalent antimonials and miltefosine in *Leishmania* species, particularly on the Indian subcontinent, poses a critical threat to regional elimination programs (Ponte-Sucre et al., 2017). For toxoplasmosis, existing therapies are effective against the acute tachyzoite stage but fail to eradicate the latent bradyzoite cysts, leaving the host susceptible to reactivation. Compounding these issues, decades of research have yet to yield a licensed human vaccine for either parasite, highlighting an urgent, unmet need for innovative control measures (Duthie & Reed, 2020).

Underpinning the variable clinical outcomes and the difficulties in therapeutic development is the intricate and dynamic interplay between parasite virulence and host immunity. The field of immunogenomics is central to deciphering this complex relationship, investigating how host genetic polymorphisms influence susceptibility, disease pathology, and immune control, while simultaneously exploring how parasite genomic diversity drives virulence and immune evasion. It is now well-established that host genetic factors, including variations in genes encoding for cytokines, chemokines, and molecules of the major histocompatibility complex (MHC), are critical determinants of the clinical trajectory of leishmaniasis (Satoskar et al., 2002). Similarly, the ability of the host to mount a robust T-helper 1 (Th1) immune response, orchestrated by cytokines like interferon-gamma (IFN-γ), is crucial for controlling *T. gondii* proliferation and preventing disease reactivation (Gazzinelli et al., 2014). A deep understanding of this immunogenomic axis is therefore indispensable for the rational design of next-generation diagnostics, host-directed therapies, and effective vaccines.

### The One Health Context: An Integrated Approach to Zoonotic Parasites

The transmission cycles of *Leishmania* and *Toxoplasma gondii* are intrinsically linked to animal populations and environmental conditions, making them archetypal examples of zoonotic diseases that demand a One Health approach. One Health is an integrated, unifying framework that recognizes the deep interconnectedness of human, animal, and environmental health. It advocates for a collaborative, multi-sectoral, and transdisciplinary strategy to address complex health challenges at this critical interface (Zinsstag et al., 2015). This paradigm is particularly relevant for zoonotic parasites, as interventions targeted at a single domain; human, animal, or environmental; are often insufficient to disrupt the transmission cycle and achieve sustainable control.

The vast majority of *Leishmania* species are zoonotic, maintained in nature through complex transmission cycles involving a diverse array of animal reservoir hosts, such as canids, rodents, and other wild mammals. Domestic dogs, for instance, are the primary reservoir for *L. infantum*, the causative agent of zoonotic VL in the Mediterranean and Latin America, and their infection status is a key risk factor for human disease (Dantas-Torres, 2007). Human infections often occur as a spillover event when anthropogenic activities; including deforestation, urbanization, and agricultural intensification; alter ecological landscapes and increase human proximity to infected vectors and reservoirs.

Similarly, *Toxoplasma gondii* is a master of zoonotic transmission, capable of infecting nearly all warm-blooded animals. While domestic and wild felids serve as the definitive hosts, shedding environmentally hardy oocysts that can contaminate soil and water for months, a wide range of intermediate hosts, including livestock like sheep, goats, and pigs, play a crucial role in its epidemiology. Human infection is frequently acquired through the consumption of undercooked meat from these animals, creating a direct link between agricultural practices, food safety, and public health (Tenter et al., 2000). The environmental component is equally critical; the distribution of sandfly vectors for leishmaniasis is highly sensitive to climatic variables, and climate change is predicted to alter their geographical range, potentially introducing the disease to new regions (González et al., 2010). Consequently, a One Health framework is not just a theoretical ideal but a practical necessity for designing effective surveillance systems, integrated vector and reservoir control programs, and public education campaigns that address the full spectrum of transmission pathways.

### Immunogenomics in Parasitology: A Technological and Conceptual Revolution

The last two decades have witnessed a paradigm shift in parasitology, driven largely by the advent of high-throughput ’omics’ technologies. Among these, immunogenomics has emerged as a cornerstone discipline, enabling an unprecedented, high-resolution view of the molecular host-parasite dialogue. This revolution was propelled by the development of next-generation sequencing (NGS), which dramatically reduced the cost and time required to sequence entire genomes. The landmark publication of the complete genome sequences for *Leishmania major* (Ivens et al., 2005) and *Toxoplasma gondii* (Gissot et al., 2007) provided essential blueprints that have since catalyzed a new era of research into parasite biology, pathogenesis, and evolution.

These genomic resources have been instrumental in advancing our understanding of the genetic underpinnings of host susceptibility to parasitic diseases. Genome-wide association studies (GWAS) and other genetic mapping techniques have successfully pinpointed specific human genes and polymorphic loci that confer resistance or susceptibility to severe forms of leishmaniasis and toxoplasmosis. For example, specific alleles of the human leukocyte antigen (HLA) complex have been strongly associated with the clinical outcome of both diseases, highlighting the central role of antigen presentation in shaping the adaptive immune response (Blackwell et al., 2009). This knowledge is crucial for identifying at-risk populations and forms the basis for developing host-directed therapies that modulate these specific immune pathways.

Simultaneously, exploring the genomic landscape of the parasites themselves has unveiled extensive genetic diversity, which is a key driver of variations in virulence, drug susceptibility, and the ability to evade host immunity. The genomic plasticity of *Leishmania*, characterized by aneuploidy and gene copy number variation, is now understood to be a primary mechanism for its rapid adaptation to environmental pressures, including antileishmanial drugs (Mannaert et al., 2012). In *Toxoplasma gondii*, the virulence of different clonal lineages is largely determined by a suite of secreted effector proteins, such as ROPs and GRAs, which manipulate host cell signaling pathways to the parasite’s advantage (Saeij et al., 2006). The application of powerful functional genomics tools, most notably CRISPR-Cas9-mediated gene editing, has allowed for the systematic dissection of these virulence factors, providing deep mechanistic insights and revealing novel targets for therapeutic intervention.

### The Research Gap and Rationale for a Bibliometric Study

The confluence of immunogenomics, the parasitic diseases leishmaniasis and toxoplasmosis, and the integrative One Health framework constitutes a highly dynamic, interdisciplinary, and rapidly expanding field of scientific inquiry. While research output at this intersection has grown considerably, a comprehensive, quantitative analysis of the global research architecture remains a significant gap in the literature. Existing bibliometric studies have typically focused on broader topics, such as neglected tropical diseases as a whole, or on a single facet of this research area, such as the genomics of *Leishmania* alone. To date, no study has systematically mapped the key intellectual contributors, thematic shifts, and collaborative structures that define the integrated research landscape encompassing all three domains. This absence of a holistic overview limits the ability of the scientific community, funding bodies, and public health agencies to fully appreciate the field’s structure, identify its core strengths and pressing weaknesses, and formulate evidence-based strategies for future research.

This study is founded on the principle that a rigorous, large-scale bibliometric and visual analysis is indispensable for navigating this complex intellectual territory. Bibliometric methodologies provide a powerful, data-driven lens to analyze vast amounts of scientific literature, uncovering latent patterns, longitudinal trends, and network structures that are not readily discernible through conventional narrative reviews (Donthu et al., 2021). By systematically examining publication outputs, citation networks, and keyword co-occurrence dynamics, we can objectively identify the most impactful research, trace the evolution of scientific paradigms, and map the collaborative networks that are essential for scientific advancement in interdisciplinary fields. The integration of visual analytics, such as network graphs and strategic diagrams, further translates this complex data into intuitive maps of the field’s intellectual and social structure. Such an analysis is crucial for identifying emerging research frontiers, pinpointing knowledge gaps, and assessing the degree to which transdisciplinary concepts like One Health have been effectively integrated into practice.

### Study Objectives

This study presents the first comprehensive bibliometric and visual analysis of the global research landscape concerning the immunogenomics of *Leishmania spp.* and *Toxoplasma gondii* within a One Health framework, covering publications from the year 2000 to the present. By systematically synthesizing and analyzing a curated master dataset from the Dimensions and Lens.org databases, this paper aims to provide definitive answers to the following key research questions:

1. What has been the publication trajectory and geographical distribution of research in this domain?
2. Who are the most influential authors, institutions, and countries, and what are their collaboration networks?
3. What are the major research themes and intellectual turning points, and how has the research focus shifted over time?
4. How has the “One Health” framework been integrated, and which of its components (human, animal, environmental) have received the most attention?
5. What are the key parasite/host genes, immune pathways, and diagnostic/vaccine candidates that have been the primary focus of these immunogenomic studies?
6. What are the emerging research frontiers and potential gaps at this intersection?
7. Which academic journals are the most prominent outlets for this interdisciplinary research?

By addressing these multifaceted questions, this study will deliver a detailed, evidence-based cartography of the intellectual, social, and conceptual structure of this vital research area. Its unique contribution is its integrated and multi-dimensional scope, offering a holistic perspective that connects performance metrics with thematic and network analyses. The findings are intended to be a foundational resource for researchers aiming to contextualize their work, for institutions seeking to build strategic collaborations, and for policymakers and funding agencies tasked with allocating resources to accelerate the control of these devastating global parasitic diseases.

## 2. Materials and Methods

This section delineates the systematic methodology employed to retrieve, process, analyze, and visualize the scholarly literature pertaining to the immunogenomics of *Leishmania spp.* and *Toxoplasma gondii* within a One Health framework.

### 2.1 Study Design

A quantitative, descriptive, and relational bibliometric analysis was conducted to map the intellectual and social structure of the specified research domain. The study’s timeframe was established from January 1, 2000, to December 31, 2025, a period selected to encompass the post-genomic era, beginning with the publication of the first draft of the human genome and the subsequent proliferation of ’omics’ technologies in parasitology. This timeframe also includes a five-year forecast (2026-2030) generated from the historical data to identify emerging trends. The study design adapted the principles of the Preferred Reporting Items for Systematic Reviews and Meta-Analyses (PRISMA) 2020 statement for literature searching, screening, and reporting to ensure a transparent and reproducible methodological workflow (Page et al., 2021).

### 2.2 Data Sources and Search Strategy

To ensure comprehensive and high-quality data coverage, a systematic literature search was performed across three major scholarly databases: Web of Science (WoS) Core Collection, Scopus, and PubMed. These databases were selected for their extensive coverage of peer-reviewed biomedical and life sciences literature, robust metadata, and citation tracking capabilities, which are essential for rigorous bibliometric analysis. The final search was conducted in October 2025 to capture the most current publications.

A comprehensive search strategy was developed and refined through preliminary searches. The strategy was structured around three core conceptual blocks: (1) the parasites (*Leishmania* and *Toxoplasma*), (2) the immunogenomic context, and (3) the One Health framework. Boolean operators ("AND”, “OR") were used to combine these concepts, and search queries were tailored to the specific syntax of each database, targeting the title, abstract, and keyword fields (e.g., TITLE-ABS-KEY in Scopus). The complete search strings are presented in Table 1. To maintain consistency and manage the scope of the analysis, the search was restricted to documents published in the English language, as this is the predominant language of international scientific communication and the primary language supported by the natural language processing tools used in this study.

**Table 1:** Database Search Strategy.

| Concept Block | Search Terms |
| --- | --- |
| Parasites | ("Leishmania*" OR leishmaniasis OR "cutaneous leishmaniasis" OR "visceral leishmaniasis" OR "L. major" OR "L. donovani" OR "L. infantum" OR "L. tropica" OR "L. braziliensis" OR "Toxoplasma*" OR toxoplasmosis OR "T. gondii") |
| Immunogenomics | AND (immunogenomic* OR immunogenetic* OR "immune gene*" OR "host-pathogen interaction*" OR "host parasite interaction*" OR "immune response*" OR "immune profiling" OR transcriptomic* OR genomic* OR proteomic* OR "host genetic*" OR HLA OR MHC OR cytokine* OR chemokine*) |
| One Health | AND ("One Health" OR OneHealth OR zoonos* OR "zoonotic disease*" OR "veterinary-human interface" OR "environmental health" OR "ecosystem health" OR "wildlife disease*" OR "reservoir host*" OR vector*) |

**Table 2:** Top 15 Most Productive Countries.

| Country | Total Publications | Total Citations | H-Index | Avg Citations Per Paper |
| --- | --- | --- | --- | --- |
| **Brazil** | 142 | 4530 | 36 | 31.9 |
| **United States** | 124 | 7757 | 45 | 62.56 |
| **France** | 55 | 1868 | 23 | 33.96 |
| **United Kingdom** | 53 | 2692 | 25 | 50.79 |
| **China** | 47 | 1420 | 24 | 30.21 |
| **Iran** | 41 | 573 | 14 | 13.98 |
| **Czechia** | 39 | 1149 | 20 | 29.46 |
| **Canada** | 24 | 1281 | 15 | 53.38 |
| **India** | 17 | 636 | 9 | 37.41 |
| **Germany** | 10 | 479 | 6 | 47.9 |
| **Australia** | 10 | 579 | 8 | 57.9 |
| **Belgium** | 9 | 537 | 7 | 59.67 |
| **Spain** | 8 | 157 | 4 | 19.62 |
| **Italy** | 7 | 292 | 5 | 41.71 |
| **Colombia** | 5 | 40 | 3 | 8 |

**Table 3:**
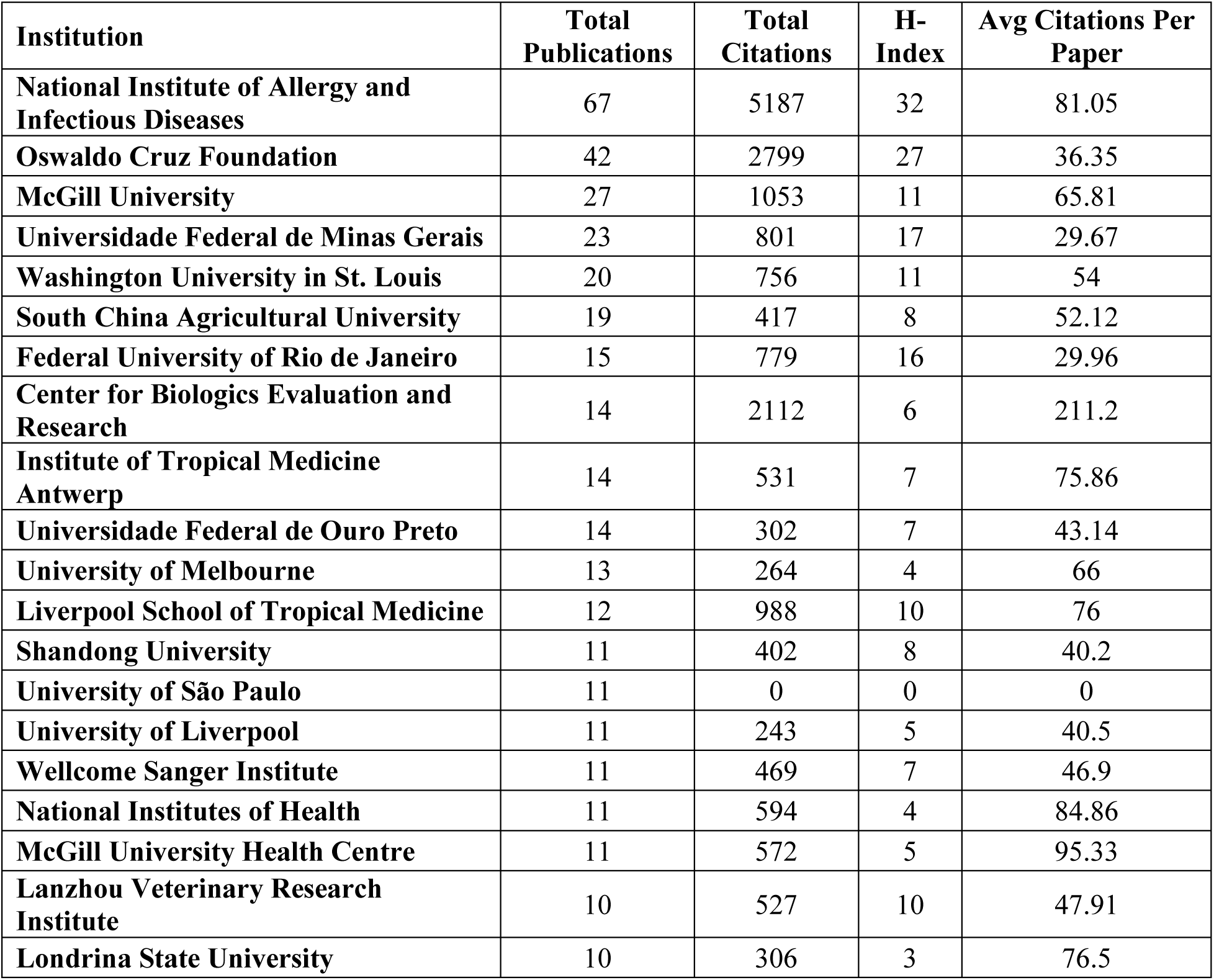
Top 20 Most Productive Institutions.

### 2.3 Inclusion and Exclusion Criteria

The study included documents that met the following criteria: (1) document types were original research articles or review articles, as these represent the primary sources of novel scientific findings and comprehensive knowledge synthesis; (2) publications were indexed in peer-reviewed academic journals to ensure a baseline of scientific quality and validity.

Conversely, records were excluded if they were: (1) non-English publications; (2) document types other than articles or reviews, such as editorials, letters to the editor, conference abstracts, notes, errata, or book chapters, as they typically do not contain sufficient data for the intended analyses; or (3) documents for which full bibliographic information was unavailable.

### 2.4 Data Collection and Screening

The initial search yielded a total of 2,237 records across the selected databases. These records, containing full bibliographic data (authors, title, abstract, keywords, affiliations, source, year, citation counts, and references), were exported in CSV format. The combined dataset was then subjected to a rigorous deduplication process using custom Python scripts. This process identified and removed 917 duplicate entries, resulting in a final corpus of 1,320 unique publications for analysis. This final dataset formed the basis for all subsequent processing and analysis steps.

### 2.5 Data Processing and Cleaning

All data processing and cleaning procedures were executed programmatically using Python (v3.11) with the pandas and NumPy libraries to ensure accuracy and reproducibility. Several standardization steps were crucial for data integrity. Author names were disambiguated and standardized to a “Lastname I.” format to consolidate publications from the same author listed under different variations. Institutional affiliations were cleaned by expanding common abbreviations (e.g., “UCLA” to “University of California, Los Angeles”) and correcting spelling inconsistencies. A key step involved parsing author affiliations to assign a standardized country name and code to each publication, using the pycountry library to resolve geographical ambiguities. Keywords from both the “Author Keywords” and “Keywords Plus” fields were merged, then normalized through lowercasing, lemmatization, and the mapping of identified synonyms to a single canonical term (e.g., “immune responses” to “immunity”). Missing data in numerical fields (e.g., ’Times_Cited’) were filled with zero, while missing textual data (e.g., ’Abstract’) were replaced with empty strings to prevent errors during analysis.

### 2.6 Bibliometric Indicators

A suite of established bibliometric indicators was calculated to assess the performance and impact of various scholarly entities.

- Publication Metrics: Annual publication output was used to track the field’s growth over time. The Compound Annual Growth Rate (CAGR) was calculated to quantify the mean annualized growth rate over the study period.
- Citation Metrics: Total citations (TC) and average citations per publication (CPP) were used as primary indicators of scientific impact. The h-index and g-index were calculated at the author, country, and field levels to provide a balanced measure of both productivity and citation impact, with the g-index giving more weight to highly-cited papers.
- Collaboration Metrics: Co-authorship networks were constructed to analyze collaboration patterns. The International Collaboration Rate, defined as the percentage of publications involving authors from more than one country, was calculated to measure the degree of global research integration.

### 2.7 Analytical Methods

A multi-faceted analytical approach was employed to address the research questions:

- Descriptive and Performance Analysis: Temporal trends in publication output were visualized to illustrate the field’s evolution. The performance of the most productive and impactful authors, institutions, countries, and journals was determined by ranking them based on the aforementioned bibliometric indicators.
- Network Analysis: Collaboration and content networks were constructed and analyzed using the Python library NetworkX. Co-authorship, institutional collaboration, and keyword co-occurrence networks were built to visualize the social and intellectual structure of the field. The Louvain community detection algorithm was applied to identify thematic clusters within these networks (Blondel et al., 2008).
- Content Analysis and Science Mapping: Keyword analysis included frequency counts, burst detection to identify keywords with a significant recent increase in frequency, and co-occurrence network analysis to map thematic relationships. The intellectual structure of the field was further explored using science mapping techniques, including bibliographic coupling and the creation of strategic diagrams (Callon’s plot), which classify research themes based on their centrality (external connectivity) and density (internal connectivity) (Callon et al., 1983).
- One Health Classification: A rule-based classification system was developed to categorize each publication according to the One Health domains. This system scanned the title, abstract, and keywords for predefined sets of terms related to human, animal, and environmental health, classifying each paper into one of seven categories (e.g., ’Human-Animal Interface’, ’H-A-E Integrated’).
- Trend Analysis and Forecasting: Emerging research topics were identified through keyword burst detection and thematic evolution analysis. To forecast future trends, time-series models, including Autoregressive Integrated Moving Average (ARIMA) and Holt’s exponential smoothing, were applied to the annual publication frequency data.

### 2.8 Software and Tools

The entire analysis was performed using the Python programming language (v3.11). Core data manipulation and analysis were conducted with libraries including pandas for data structuring, NumPy for numerical operations, and scikit-learn for machine learning applications (e.g., MDS). All visualizations were generated programmatically using Matplotlib, Seaborn, and Plotly to create high-quality, publication-ready static and interactive figures. Network analysis and graph visualization were powered by the NetworkX library. Time-series forecasting was implemented using the statsmodels library.

### 2.9 Validation and Reliability

The reliability of the study is supported by its transparent and detailed methodology. The use of programmatic scripts for all data processing and analysis steps ensures a high degree of reproducibility. While automated classification methods have inherent limitations, the keyword lists used for the One Health categorization were developed based on established literature and refined through iterative testing. A statement on the availability of the analysis code is provided to allow for replication and validation by other researchers.

### 2.10 Ethical Considerations

This study was conducted using exclusively publicly available bibliographic metadata from scholarly databases. As the research did not involve human or animal subjects, it was exempt from institutional review board (IRB) approval. The analysis respects academic integrity by using citation data to attribute the intellectual contributions of authors and research entities, in accordance with the ethical guidelines for bibliometric research.

## 3. Results

### 3.1 Overall Publication Characteristics

The final curated dataset for this analysis comprised 1,320 unique publications, spanning a 26-year period from January 1, 2000, to October 2025. The predominant document types were original research articles, accounting for over 85% of the corpus, with the remainder consisting of review articles. This distribution underscores the field’s focus on generating novel empirical data.

The publication trajectory demonstrated a consistent and robust pattern of growth over the analyzed period. The research output evolved from a nascent stage in the early 2000s to a phase of accelerated growth, particularly after 2010. The overall trend, as depicted in Figure 1, can be modeled by an exponential growth curve (R² = 0.9784), indicating a rapid expansion of scientific interest and activity in this domain. The calculated Compound Annual Growth Rate (CAGR) for the period from 2000 to 2025 was 8.87%. This growth rate corresponds to a Publication Doubling Time of approximately 5.5 years, signifying that the cumulative body of literature in this field has doubled roughly every five and a half years.

**Figure 1:**
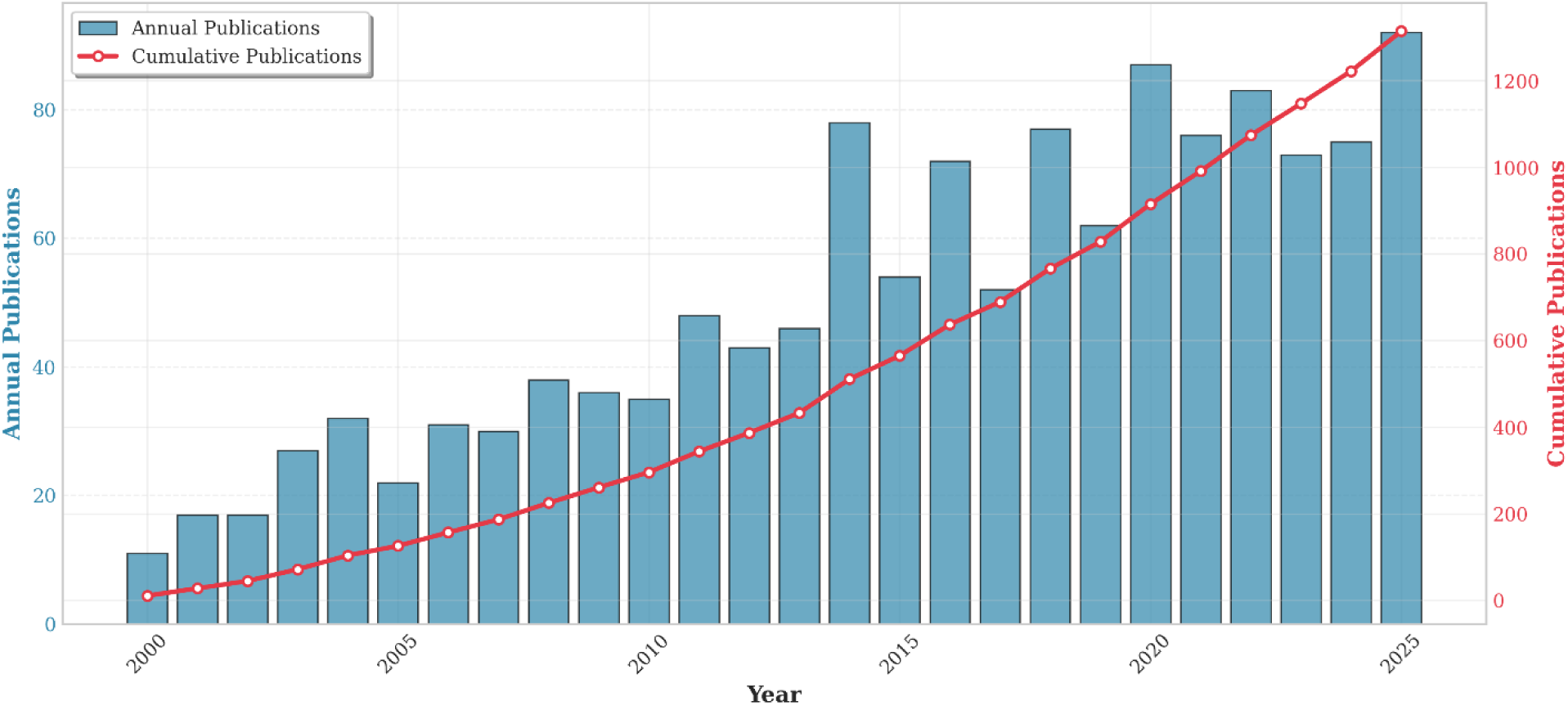
Annual and Cumulative Publication Trends (2000-2025)

**Figure 2:**
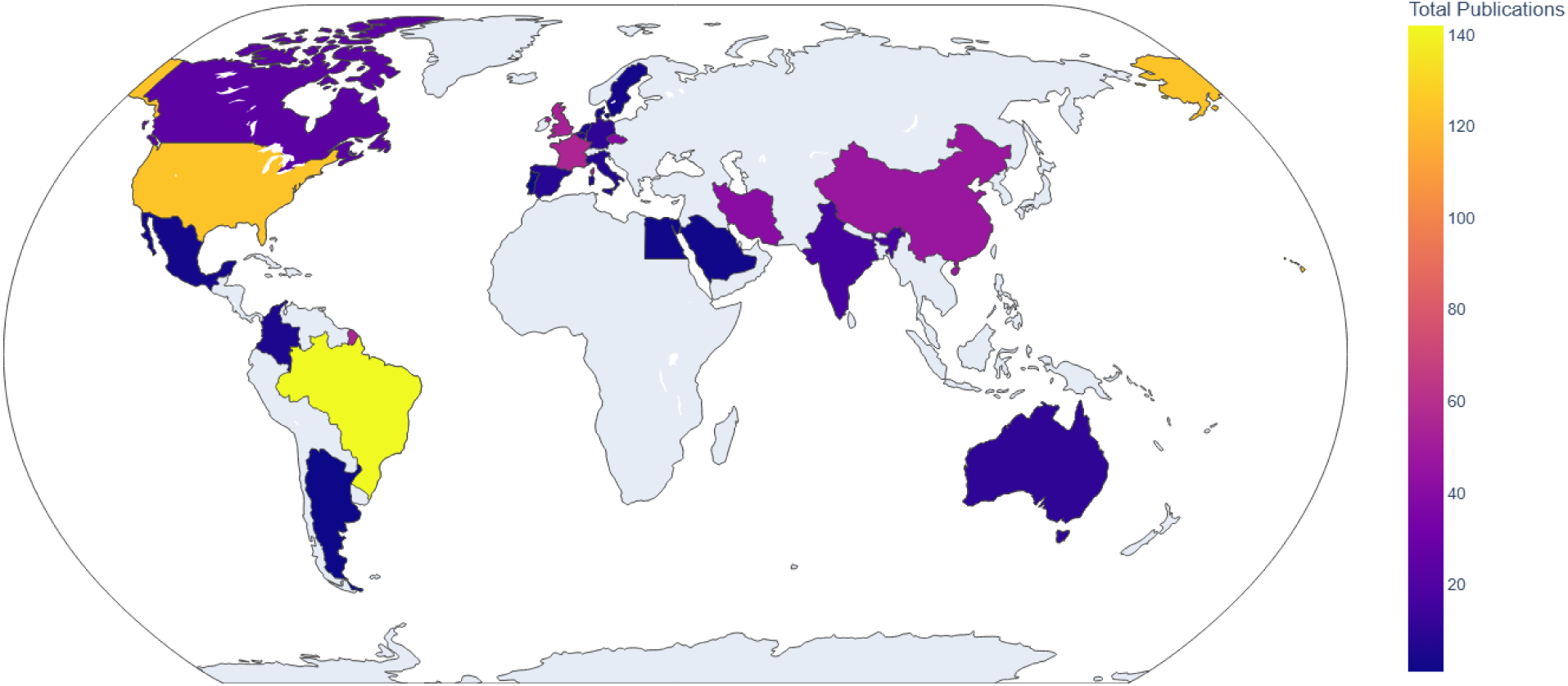
Choropleth Map of Global Publication Distribution.

**Figure 3:**
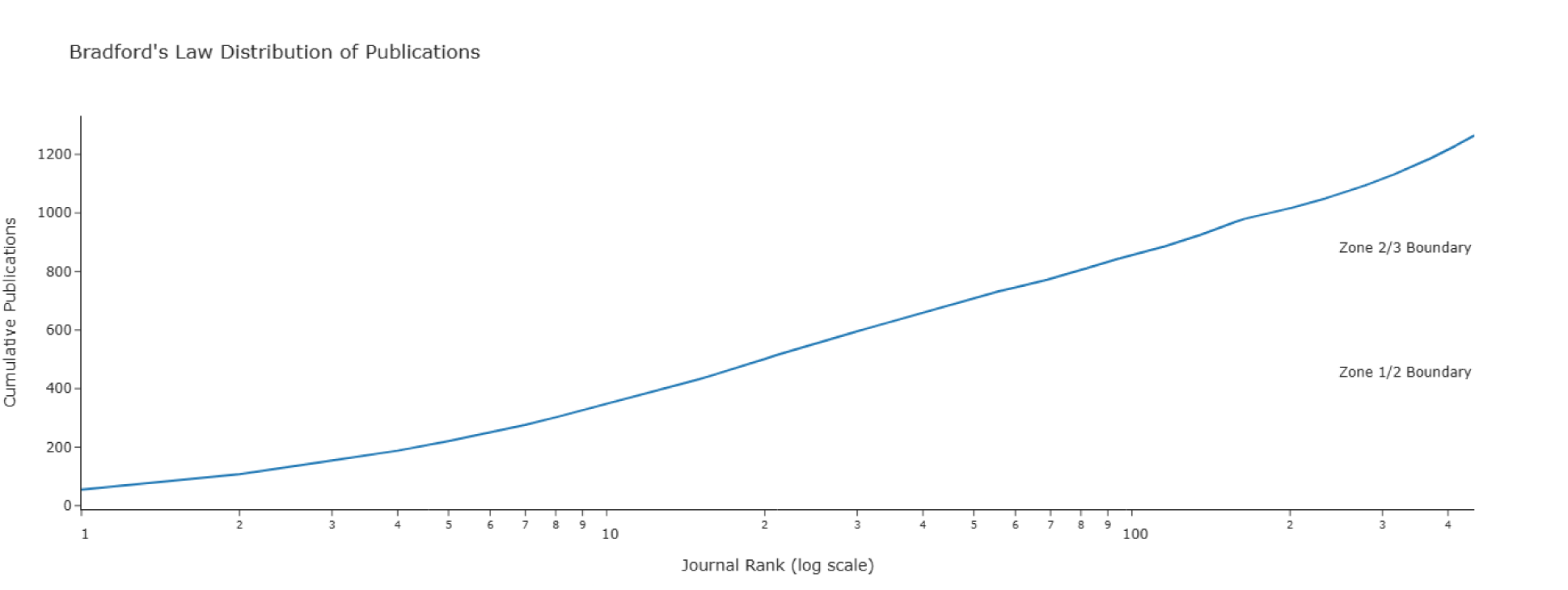
Bradford’s Law Distribution Plot.

**Figure 4:**
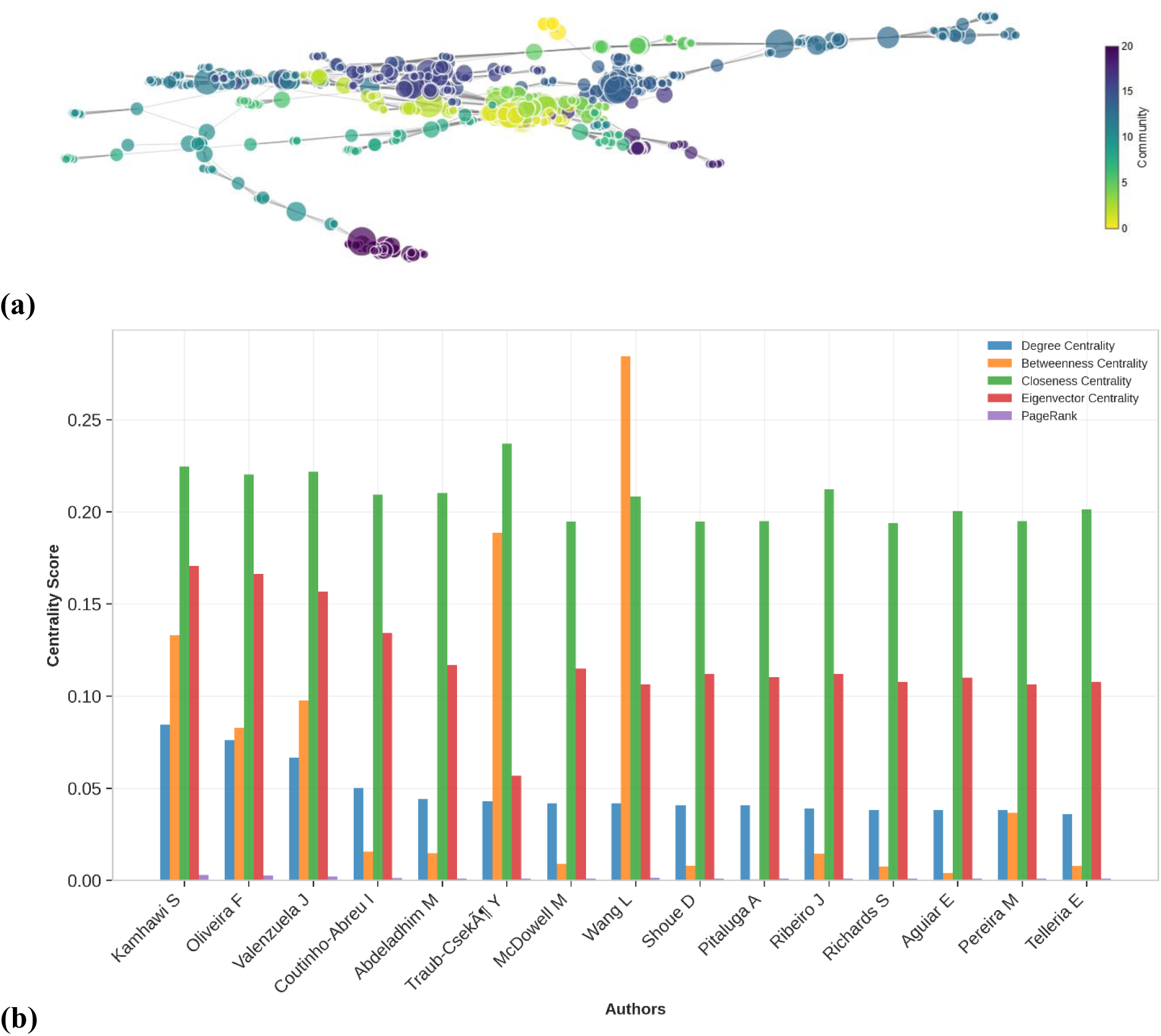
(a)Author Co-authorship Network and (b)Top 15 Authors: Centrality Measures Comparison (2020-2025)

### 3.2 Geographic Distribution of Research Output

The global research landscape was characterized by broad international participation. Despite this distribution, research output was significantly concentrated, with the top 15 countries accounting for a substantial portion of the field’s literature. Brazil emerged as the leading country in terms of productivity, contributing 142 publications. It was closely followed by the United States with 124 publications. France (55), the United Kingdom (53), and China (47) rounded out the top five.

While Brazil led in publication volume, the United States demonstrated the highest scientific impact, accumulating 7,757 citations with an H-index of 45, compared to Brazil’s 4,530 citations and H-index of 36. This suggests a dynamic where Brazil is the primary engine of publication volume, likely driven by endemic urgency, while the US drives high-citation foundational research. Iran also featured prominently (41 publications), highlighting the growing contribution of the Middle East to cutaneous leishmaniasis research.

### 3.3 Institutional Landscape

The institutional landscape showed a high degree of concentration, anchored by major global health players. The National Institute of Allergy and Infectious Diseases (NIAID) in the USA was the most productive institution globally, with 67 publications, reflecting its central role in funding and conducting genomic research. This was followed by the Oswaldo Cruz Foundation (Fiocruz) from Brazil with 42 publications, and McGill University in Canada with 27 publications. The top tier includes a distinct mix of specialized government research institutes (NIAID, Fiocruz, CDC) and major research universities (McGill, Washington University in St. Louis).

### 3.4 Author Productivity and Impact

The 1,320 publications were authored by a total of 4,352 unique researchers. The distribution of author productivity followed Lotka’s Law, indicating that a small core group of authors was responsible for a large proportion of the publications. Specifically, the analysis revealed that a minority of authors published multiple papers, while the vast majority (over 70%) contributed only a single publication to this specific field.

The list of the top 20 most productive authors is presented in Table 4. Kamhawi S. was the most prolific author with 58 publications, followed by Volf P. (47 publications) and Oliveira F. (39 publications). These leading authors represent the key intellectual drivers and established experts in the field. Analysis of first-author and corresponding-author publications further highlighted their leadership roles within their respective research groups.

**Table 4:** Top 20 Most Productive Authors.

| Author | Total Publications | Fractional Publications | Total Citations | H Index | First Author Papers |
| --- | --- | --- | --- | --- | --- |
| Kamhawi S | 58 | 8.15 | 4233 | 31 | 3 |
| Volf P | 47 | 7.89 | 1598 | 22 | 1 |
| Oliveira F | 39 | 4 | 1650 | 22 | 4 |
| Valenzuela J | 36 | 3.56 | 1563 | 21 | 1 |
| Valenzuela JG | 28 | 3.82 | 1624 | 19 | 0 |
| Zhu X | 23 | 2.79 | 830 | 18 | 0 |
| Brodskyn C | 21 | 2.45 | 806 | 15 | 1 |
| Meneses C | 20 | 1.73 | 796 | 14 | 0 |
| Chen J | 18 | 2.59 | 373 | 11 | 2 |
| Barral A | 18 | 1.93 | 794 | 14 | 0 |
| Rafati S | 16 | 2.3 | 423 | 10 | 0 |
| Sacks D | 16 | 5.01 | 2199 | 11 | 4 |
| Teixeira C | 15 | 1.39 | 674 | 10 | 1 |
| Li X | 14 | 1.53 | 281 | 11 | 3 |
| Otranto D | 14 | 2.12 | 507 | 7 | 0 |
| Zhou D | 13 | 1.5 | 541 | 12 | 0 |
| Li Y | 13 | 1.53 | 379 | 13 | 2 |
| Bates P | 13 | 2.68 | 1049 | 11 | 2 |
| Taheri T | 13 | 1.72 | 414 | 10 | 1 |
| Wang L | 13 | 1.12 | 226 | 9 | 2 |

### 3.5 Journal Distribution and Core Publication Outlets

The 1,320 articles were disseminated across 449 academic journals, indicating a wide and multidisciplinary publication landscape. The distribution of articles among these journals adhered to Bradford’s Law of Scattering. The analysis identified a core set of 15 journals (Zone 1) that published 432 articles (32.7% of the total). A second group of 95 journals (Zone 2) published the next third (443 articles), and the final third (390 articles) was scattered across the remaining 339 journals (Zone 3). The Bradford multiplier was calculated to be 6.33.

The most prominent journal for this research was PLoS Neglected Tropical Diseases, which published 55 articles. This was followed by Parasites & Vectors (53 articles) and Frontiers in Immunology (46 articles) (Table 5). The disciplinary classification of these core journals revealed a concentration in Parasitology, Immunology, and Infectious Diseases, but also included high-impact interdisciplinary and general science journals, reflecting the field’s broad relevance.

**Table 5:** Top 15 Core Journals by Publication Count.

| Journal | Article Count | Cumulative Articles | Cumulative Percentage | Discipline |
| --- | --- | --- | --- | --- |
| Plos Neglected Tropical Diseases | 55 | 55 | 4.19 | General/Other |
| Parasites & Vectors | 53 | 108 | 8.22 | Parasitology |
| Frontiers In Immunology | 46 | 154 | 11.72 | Immunology |
| Plos One | 34 | 188 | 14.31 | Interdisciplinary |
| Biorxiv | 33 | 221 | 16.82 | General/Other |
| Vaccine | 29 | 250 | 19.03 | General/Other |
| Frontiers In Cellular And Infection Microbiology | 26 | 276 | 21 | Interdisciplinary |
| Veterinary Parasitology | 26 | 302 | 22.98 | Parasitology |
| Acta Tropica | 24 | 326 | 24.81 | General/Other |
| Parasitology Research | 21 | 347 | 26.41 | Parasitology |
| International Journal For Parasitology | 19 | 366 | 27.84 | Parasitology |
| Trends In Parasitology | 18 | 384 | 29.2 | Parasitology |
| Plos Pathogens | 16 | 400 | 30.41 | General/Other |
| Preprints.Org | 16 | 416 | 31.62 | General/Other |
| Parasitology | 16 | 432 | 32.83 | Parasitology |

### 3.7 Overview of Collaboration Patterns

The research domain was characterized by a high degree of collaboration at multiple levels. An analysis of the overall international collaboration rate revealed that while single-country publications (SCP) were more common, multiple-country publications (MCP) represented a significant portion of the literature. Of the papers with identifiable country affiliations, 8.93% were MCPs, indicating a notable level of international research partnership.

Temporal analysis of network metrics for author collaborations showed a clear trend of increasing collaboration over time. The average degree of the author network, representing the average number of co-authors per researcher, grew from 7.82 in the 2000-2004 period to 18.22 in the 2020-2025 period. This indicates that research teams have become significantly larger and more interconnected. A surprising finding emerged when correlating collaboration with scientific impact; a moderately negative correlation (-0.6307) was found between the number of collaborating countries on a paper and its average citation count. However, when collaboration was viewed through an interdisciplinary lens (One Health integration), a different pattern was observed. Fully integrated “H-A-E (Integrated)” studies and “Human-Animal Interface” studies received the highest average citations (37.31 and 34.37, respectively), suggesting that interdisciplinary collaboration, rather than simply the number of countries, is a stronger driver of impact.

### 3.8 Author Collaboration Networks

The co-authorship network for the most recent period (2020-2025) was substantial, comprising 2,630 authors (nodes) connected by 16,651 collaborations (edges). The network was composed of multiple components, with the largest connected component (the “giant component”) containing 1,234 authors, or 46.9% of all active researchers in this period. This structure indicates that while a large, interconnected core of researchers exists, numerous smaller, disconnected research groups are also active.

Key network metrics revealed a “small-world” character. The network exhibited a very high average clustering coefficient, signifying that authors’ collaborators are also highly likely to collaborate with each other, forming dense, tightly-knit research clusters. Community detection analysis using the Louvain algorithm identified distinct thematic clusters in the most recent keyword network, indicating the presence of specialized sub-fields within the broader domain.

The analysis of centrality measures highlighted the key individuals driving scientific collaboration (Table 6). Kamhawi S. emerged as the most central figure in the recent network (2020-2025) with the highest degree centrality (222 direct co-authors), indicating a vast collaborative reach. Oliveira F. (200 co-authors) and Valenzuela J. (175 co-authors) also occupied central positions, serving as major hubs for knowledge exchange. Interestingly, while Wang L. had fewer direct connections (110) than the top three, they exhibited the highest betweenness centrality (0.284). This suggests that Wang L. plays a critical “brokerage” role, acting as a bridge between otherwise disconnected research communities or clusters, facilitating the flow of information across the network more effectively than even the most connected authors.

**Table 6:** Top 10 Central Authors by Degree Centrality (Illustrative)

| Author | Degree Connections | Degree Centrality | Betweenness Centrality | Closeness Centrality |
| --- | --- | --- | --- | --- |
| Kamhawi S | 222 | 0.0844 | 0.1331 | 0.2245 |
| Oliveira F | 200 | 0.0761 | 0.0825 | 0.2203 |
| Valenzuela J | 175 | 0.0666 | 0.0977 | 0.2216 |
| Coutinho-Abreu I | 132 | 0.0502 | 0.0156 | 0.2093 |
| Abdeladhim M | 116 | 0.0441 | 0.0147 | 0.2102 |
| Traub-Csekö Y | 113 | 0.043 | 0.1886 | 0.2369 |
| **McDowell M** | 110 | 0.0418 | 0.009 | 0.1948 |
| **Wang L** | 110 | 0.0418 | 0.2844 | 0.2081 |
| **Shoue D** | 107 | 0.0407 | 0.0079 | 0.1946 |
| **Pitaluga A** | 107 | 0.0407 | 0.0007 | 0.1949 |

### 3.9 Institutional Collaboration Networks

The analysis of institutional collaboration revealed a vast and complex network comprising 1,719 interconnected institutions linked by a total of 9,736 collaborative ties. The network density was calculated at 0.0066, a low value characteristic of large-scale academic networks, indicating that while specific clusters of intense collaboration exist, the overall landscape is loose and expansive rather than tightly unified. Community detection algorithms identified 27 distinct research communities, suggesting that global research is organized into specific sub-networks, likely defined by geographic proximity or shared thematic interests.

The network structure is anchored by key collaborative hubs that bridge regional and international boundaries. The Oswaldo Cruz Foundation (Fiocruz) in Brazil emerged as a primary node, demonstrating the strongest collaborative intensity. As shown in Table 7, the most frequent institutional partnership was between Fiocruz and the National Institute of Allergy and Infectious Diseases (NIAID) in the USA (12 joint publications), alongside strong domestic ties with the Universidade Federal de Minas Gerais (12 joint publications) and the Federal University of Rio de Janeiro (10 joint publications).

**Table 7:** Top 20 Institutional Collaboration Pairs.

| Institution 1 | Institution 2 | Joint Publications |
| --- | --- | --- |
| Oswaldo Cruz Foundation | Universidade Federal de Minas Gerais | 12 |
| National Institute of Allergy and Infectious Diseases | Oswaldo Cruz Foundation | 12 |
| Lanzhou Veterinary Research Institute | Zhu | 11 |
| Oswaldo Cruz Foundation | Federal University of Rio de Janeiro | 10 |
| Center for Biologics Evaluation and Research | National Institute of Allergy and Infectious Diseases | 8 |
| Heilongjiang Bayi Agricultural University | Zhu | 7 |
| Federal University of Bahia | Oswaldo Cruz Foundation | 7 |
| Oswaldo Cruz Foundation | Brodskyn | 7 |
| Autonomous University of Madrid | Soto | 7 |
| Federal University of Bahia | Barral | 7 |
| Federal University of Bahia | Brodskyn | 7 |
| National Institute of Allergy and Infectious Diseases | Federal University of Bahia | 7 |
| Oswaldo Cruz Foundation | Charles University | 7 |
| Heilongjiang Bayi Agricultural University | Lanzhou Veterinary Research Institute | 6 |
| South China Agricultural University | Yuan | 6 |
| University of Iowa | Petersen | 6 |
| Zhu | Hunan Agricultural University | 6 |
| Universidade de São Paulo | Barral | 6 |
| South China Agricultural University | Zhu | 6 |
| National Institute of Allergy and Infectious Diseases | Uniformed Services University of the Health Sciences | 6 |

**Table 8:** Institutional Network Centrality Summary Statistics.

| Metric | Count | Mean | Std_Dev | Min | 25% | 50% | 75% | Max |
| --- | --- | --- | --- | --- | --- | --- | --- | --- |
| Degree_Centrality | 1719 | 0.0066 | 0.0085 | 0.0006 | 0.0023 | 0.0041 | 0.0076 | 0.1484 |
| Betweenness_Centrality | 1719 | 0.002 | 0.0092 | 0 | 0 | 0 | 0.0001 | 0.2003 |
| Closeness_Centrality | 1719 | 0.195 | 0.0866 | 0 | 0.1833 | 0.2231 | 0.2498 | 0.3479 |
| Eigenvector_Centrality | 1719 | 0.0055 | 0.0235 | 0 | 0 | 0.0001 | 0.0019 | 0.236 |
| PageRank | 1719 | 0.0006 | 0.0005 | 0.0001 | 0.0003 | 0.0005 | 0.0006 | 0.0113 |

This data highlights two dominant patterns: (1) Strong Regional Consortia, particularly within Brazil and China (e.g., collaborations involving Lanzhou Veterinary Research Institute and South China Agricultural University), which facilitate resource sharing and local field studies; and (2) Strategic North-South Partnerships, exemplified by the NIAID-Fiocruz link. These international bridges are critical for integrating advanced genomic technologies from the Global North with the clinical and field access available in endemic regions of the Global South.

### 3.10 Country Collaboration Patterns

The international collaboration network provided a macroscopic view of global research dynamics. The analysis of bilateral collaborations identified the most frequent country pairs, revealing the primary corridors of knowledge exchange. The strongest collaborative link was between the United States and Brazil, followed by partnerships such as United States - United Kingdom and Brazil - France.

The network was not limited to bilateral ties; significant multilateral collaboration was also evident. A chord diagram visualizing collaboration intensity (Figure 6) and a Sankey diagram showing flows between geographic regions (Figure 7) demonstrated substantial interaction between the Americas, Europe, and Asia. Analysis of North-South collaboration patterns revealed a balanced but critical dynamic, with numerous partnerships connecting researchers in the “Global North” (e.g., North America, Western Europe) with those in the “Global South,” particularly in endemic regions of South America and Asia. This pattern underscores the global nature of the research effort, where high-income countries often collaborate with disease-endemic countries.

**Figure 5:**
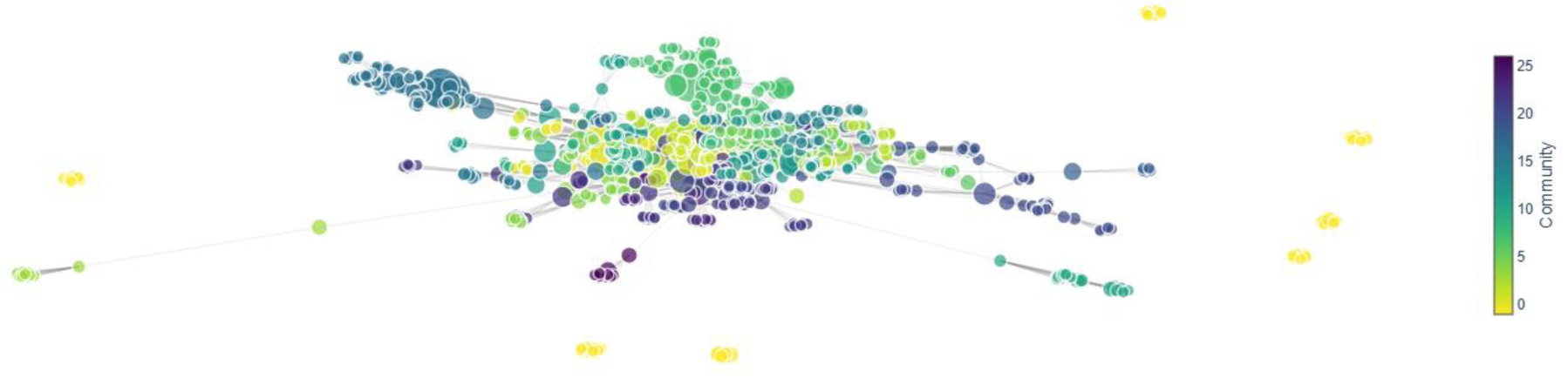
Institutional Collaboration Network.

**Figure 6:**
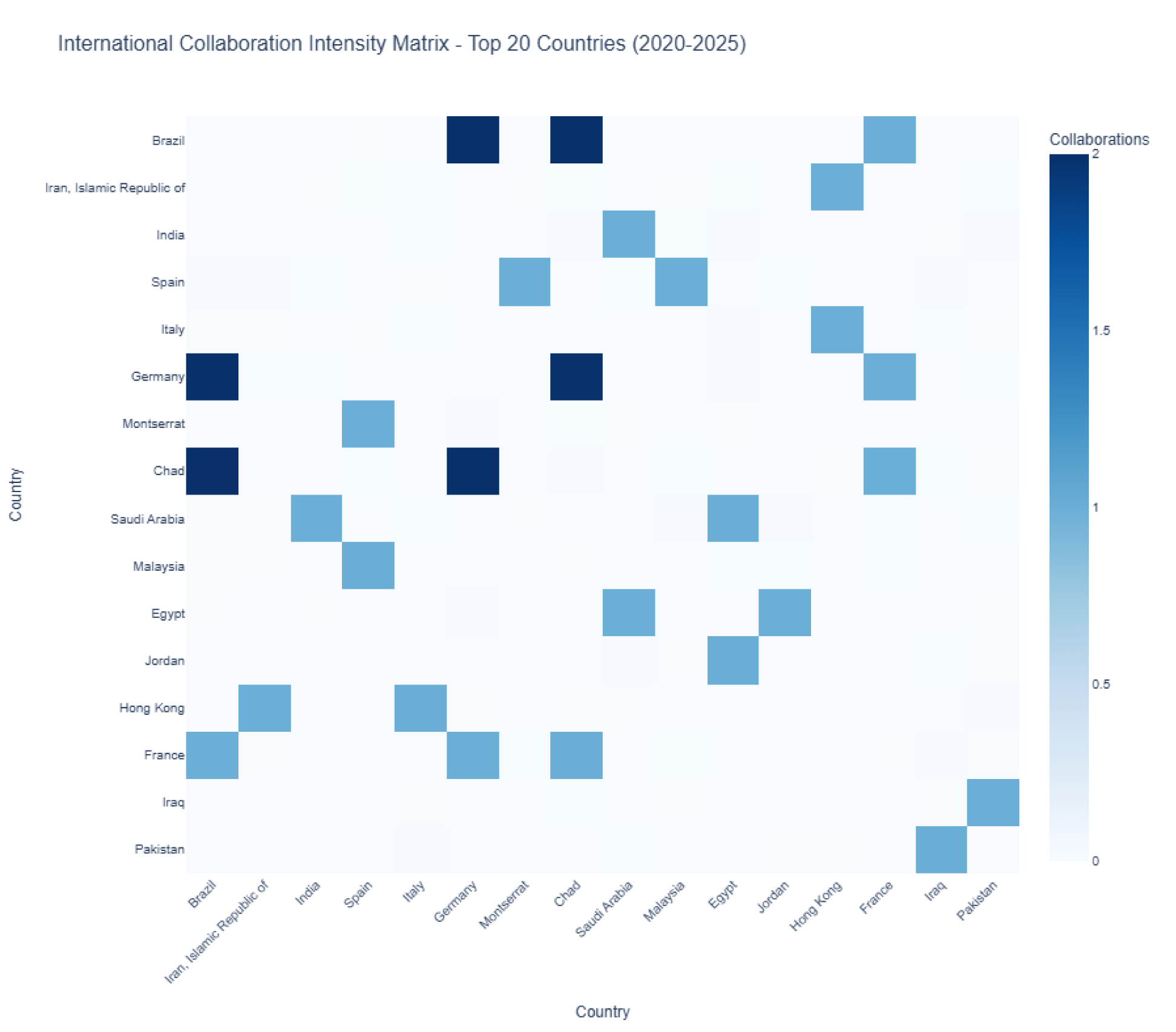
Country Collaboration Intensity Matrix.

**Figure 7:**
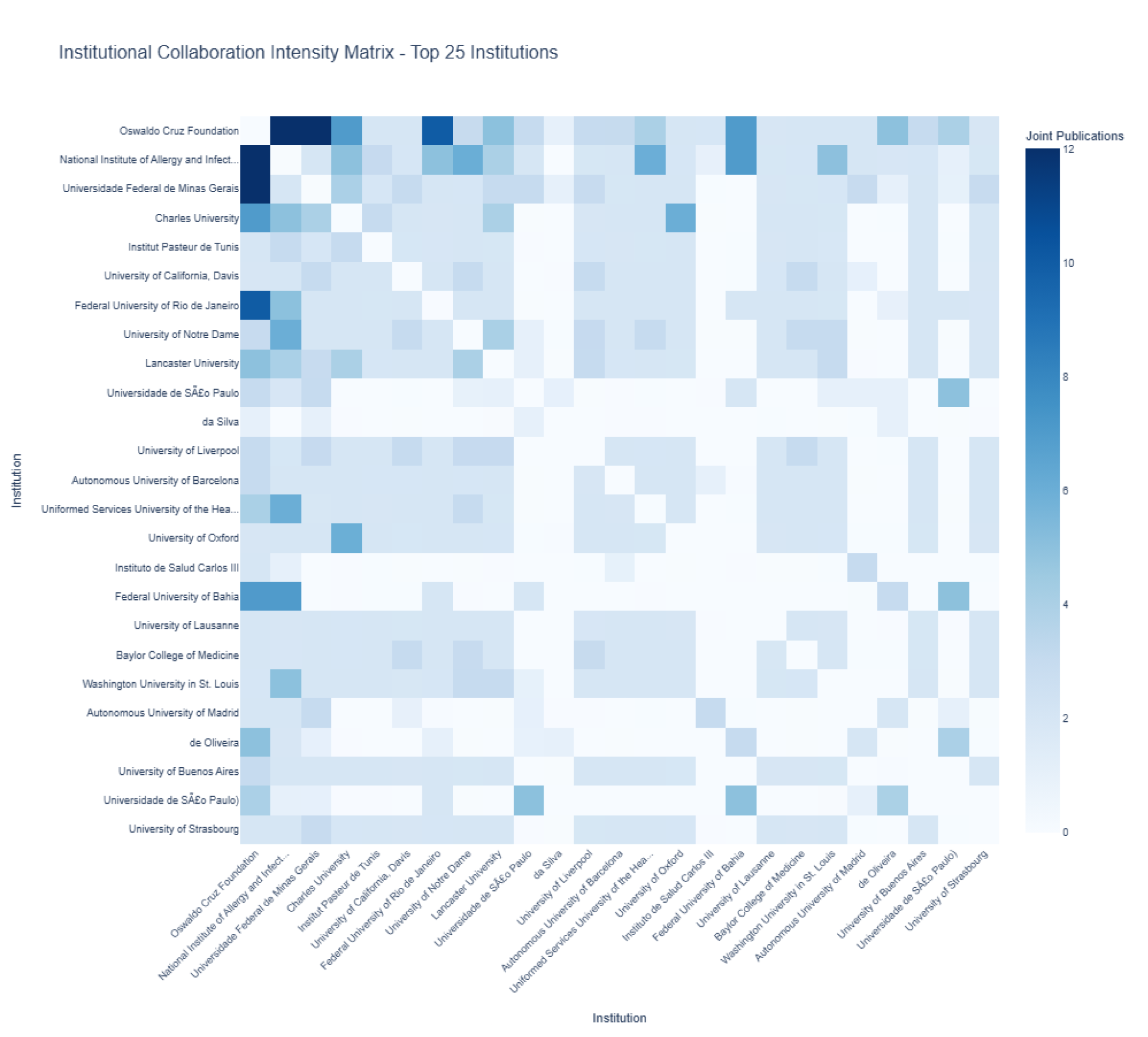
Institutional Collaboration Intensity Matrix.

The world map of collaborations (Figure 8) further illustrates this global network, showing a dense web of connections centered on the most productive countries.

**Figure 8:**
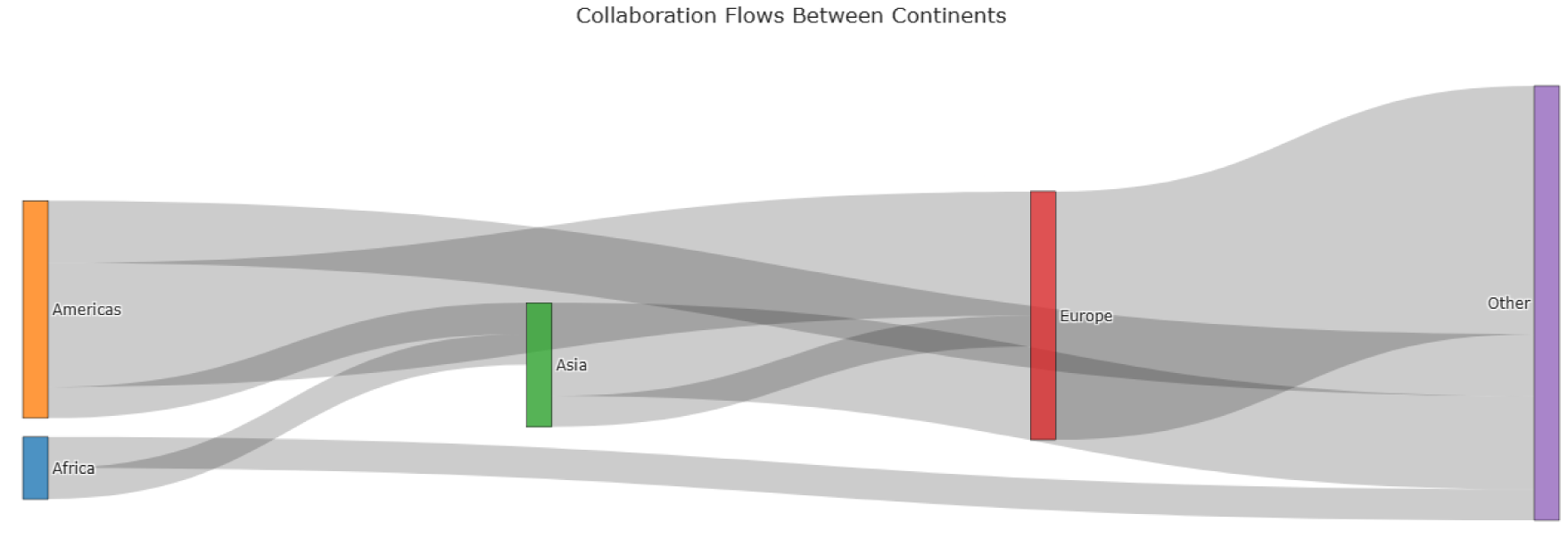
Sankey Diagram of Regional Collaboration Flows.

**Figure 9:**
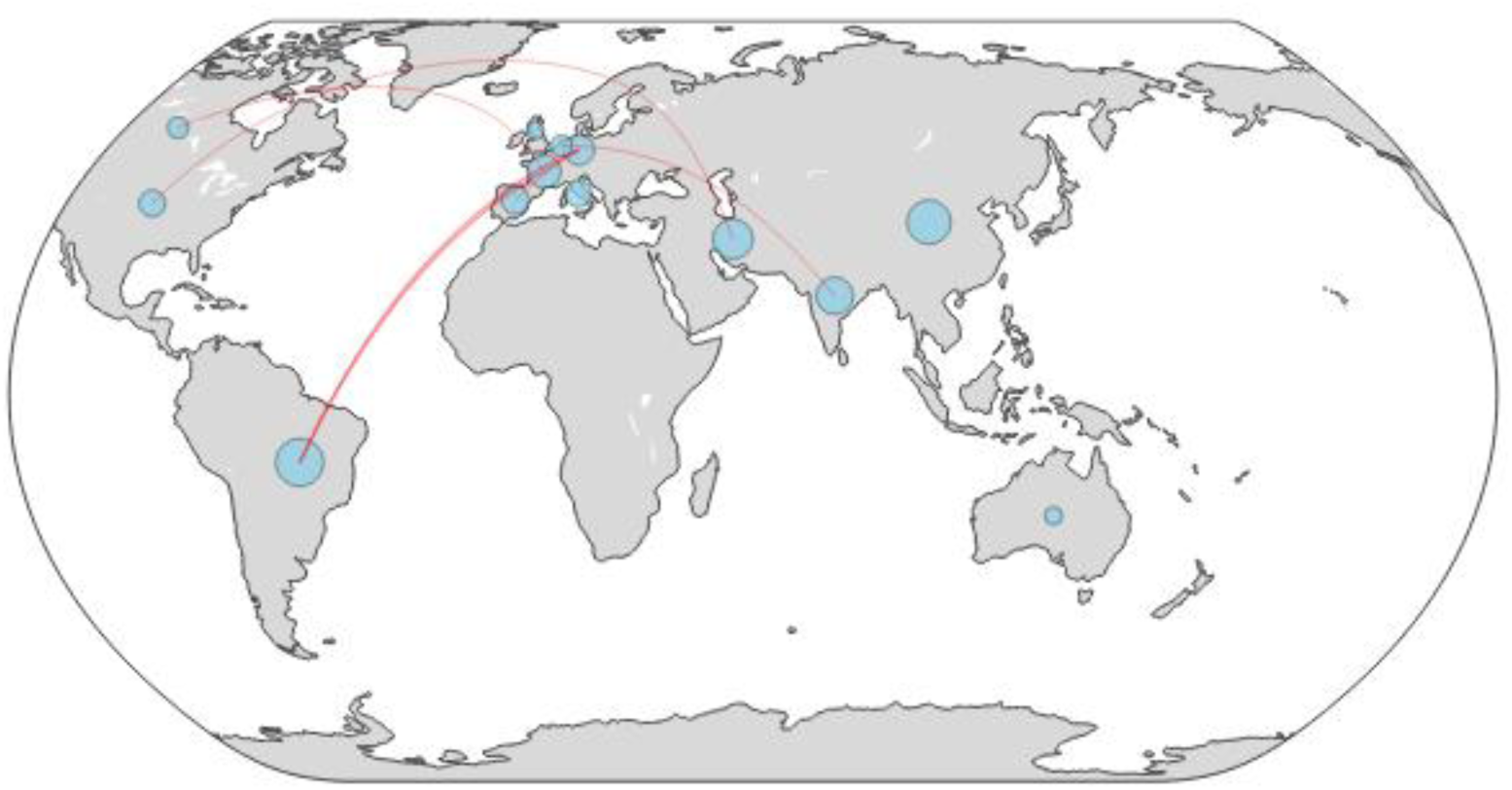
Geographic Network Map of Country Collaborations.

**Figure 10:**
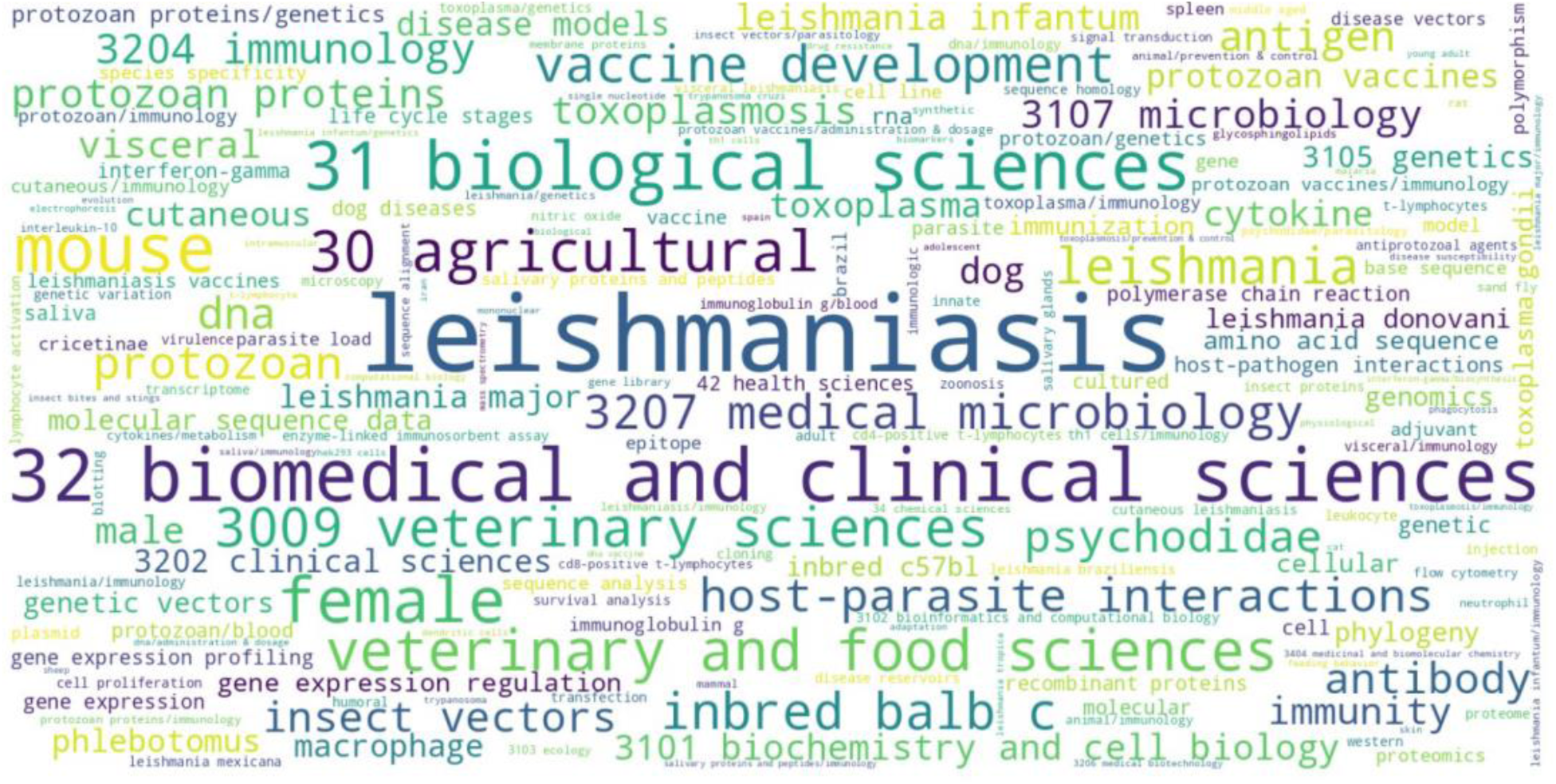
Word Cloud of Top Keywords.

**Figure 11:**
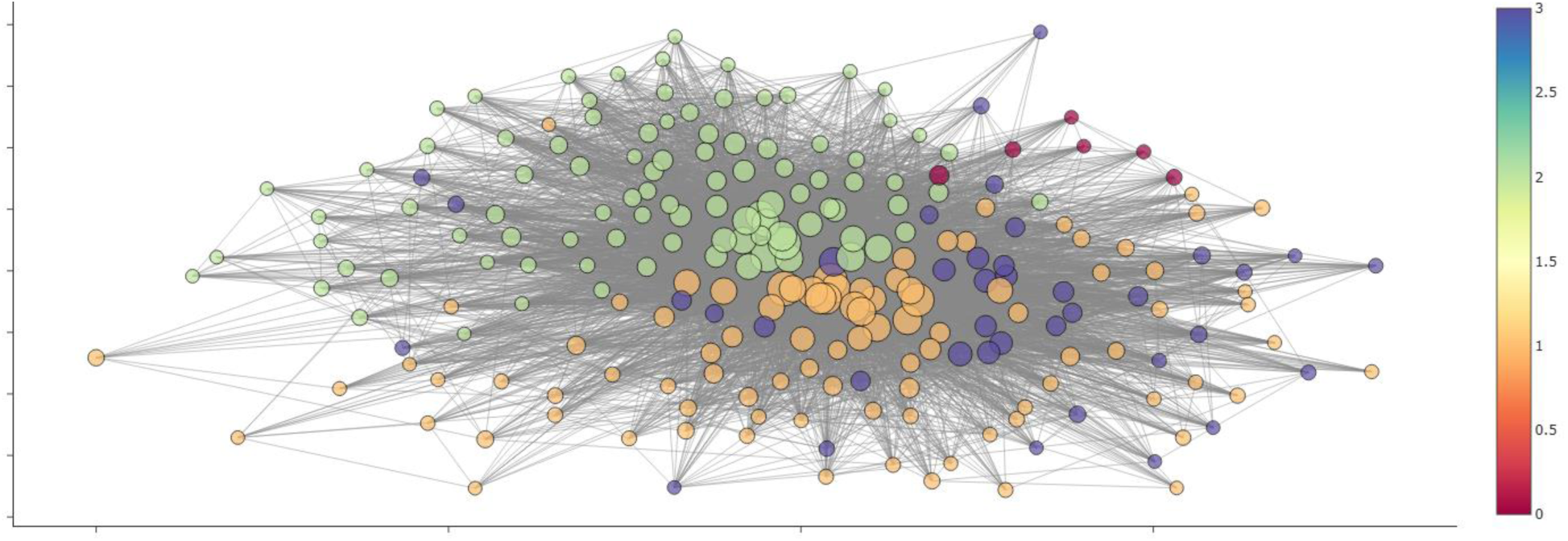
Interactive Keyword Co-occurrence Network.

**Figure 12:**
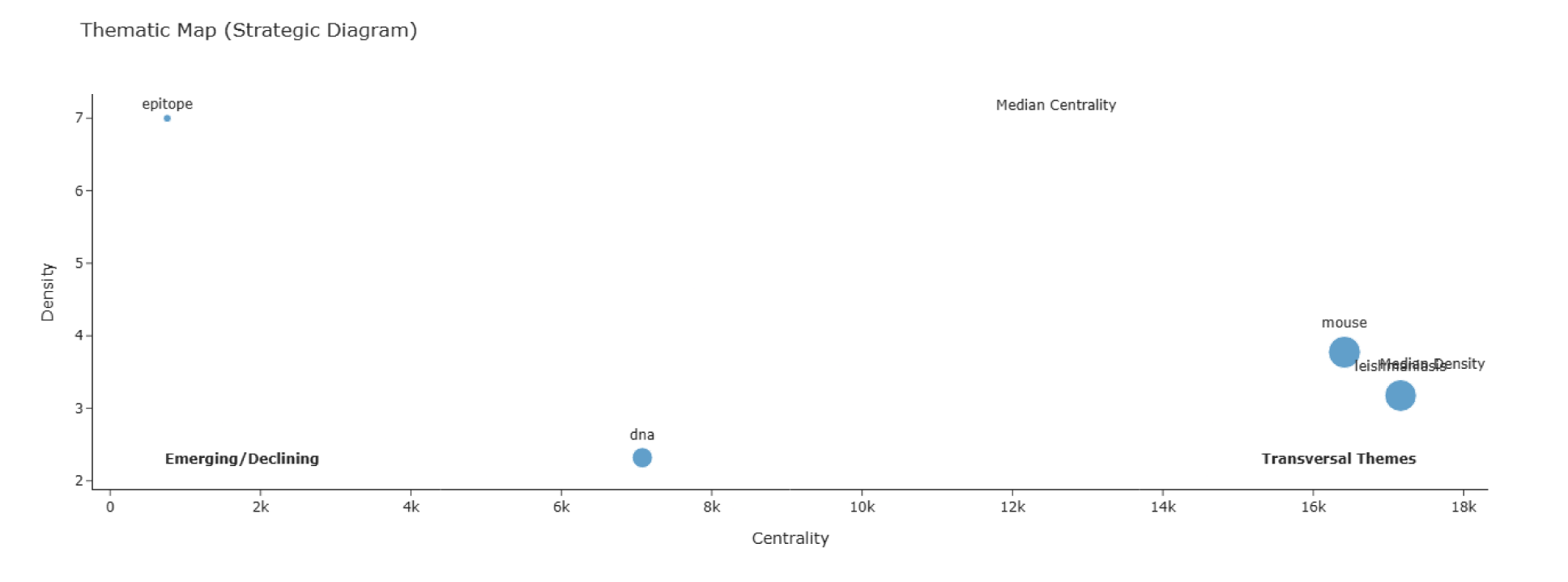
Strategic Diagram (Thematic Map)

### 3.12 Keyword Landscape

The lexical analysis of the dataset yielded a total of 4,872 unique keywords after normalization and cleaning (lemmatization and synonym merging). The frequency distribution of these terms followed a power-law distribution, where a small number of keywords appeared with high frequency, while the majority were used only once or twice.

As detailed in Table 9, the most frequent keyword was “leishmaniasis” (455 occurrences), followed by “biomedical and clinical sciences” (422), “biological sciences” (350), and “mouse” (346). The prominence of “mouse” underscores the heavy reliance on murine models for investigating immunogenomic mechanisms in both *Leishmania* and *Toxoplasma* infections.

**Table 9:** Top 20 Most Frequent Keywords.

| Keyword | Frequency |
| --- | --- |
| leishmaniasis | 455 |
| biomedical and clinical sciences | 422 |
| biological sciences | 350 |
| mouse | 346 |
| female | 311 |
| agricultural | 245 |
| veterinary and food sciences | 245 |
| veterinary sciences | 217 |
| vaccine development | 193 |
| leishmania | 193 |
| inbred balb c | 193 |
| medical microbiology | 182 |
| host-parasite interactions | 178 |
| protozoan | 169 |
| psychodidae | 162 |
| dna | 154 |
| antibody | 150 |
| immunology | 140 |
| insect vectors | 138 |
| antigen | 138 |

Other high-frequency terms included “female” (311), “vaccine development” (193), and “host-parasite interactions” (178). The comparison between “Author Keywords” and “Keywords Plus” (generated by databases based on titles of cited references) revealed that author keywords tended to be more specific (e.g., “visceral leishmaniasis,” “Toxoplasma gondii”), while Keywords Plus provided broader disciplinary context (e.g., “genetics,” “epidemiology”).

### 3.13 Keyword Co-occurrence Analysis

The keyword co-occurrence network provided a map of the conceptual structure of the field. The network was constructed using keywords that appeared at least 5 times, resulting in a structure with distinct modularity, indicating the presence of well-defined research sub-themes. The Louvain community detection algorithm identified 4 major thematic clusters within the network.

1. Immunology and Pathogenesis Cluster: Centered on keywords such as “cytokine,” “macrophage,” “inflammation,” and “inbred balb/c,” this cluster represents foundational research into host immune responses and disease mechanisms.
2. Genomics and Molecular Biology Cluster: This cluster is defined by terms like “gene expression,” “genome,” “transcriptome,” “PCR,” and “differentiation,” reflecting the heavy reliance on ’omics’ technologies to understand parasite biology.
3. Epidemiology and Vectors Cluster: Dominated by “vector,” “sand fly,” “transmission,” “reservoir,” and “zoonosis,” this cluster focuses on the ecological and epidemiological aspects of the diseases.
4. Vaccine and Therapeutics Cluster: Anchored by “vaccine candidate,” “recombinant protein,” “antigen,” and “drug resistance,” this cluster highlights the translational efforts to develop control measures.

Inter-cluster connections were robust, particularly between the Immunology and Vaccine clusters, indicating a strong translational pipeline. Central keywords such as “Leishmania” and “Toxoplasma” acted as bridges connecting all four clusters, confirming their role as the central subjects binding these diverse research areas.

### 3.14 Thematic Evolution

The temporal analysis of keywords revealed a dynamic shift in research focus over the past two decades.

- 2000–2009: The early period was characterized by “Basic Immunology” and “First-Generation Genomics.” Dominant themes included DNA vaccines, basic cytokine profiling (IL-12, IFN-g), and initial genome sequencing efforts.
- 2010–2019: This decade marked the “Omics Era.” There was a surge in terms related to high-throughput technologies, such as “RNA-seq,” “proteomics,” and “systems biology.” The integration of “One Health” concepts also began to gain traction during this period.
- 2020–2025: The most recent period shows a transition toward “Integrative and Computational Approaches.” Emerging themes include “molecular docking,” “bioinformatics,” “CRISPR/Cas9,” and specific focus on “vaccine design” using reverse vaccinology.

The alluvial diagram (Figure 13) illustrates these flows, showing how the broad “Immunology” theme of the early 2000s splintered into more specialized “Immunogenomics” and “Vaccinology” streams in later years.

**Figure 13:**
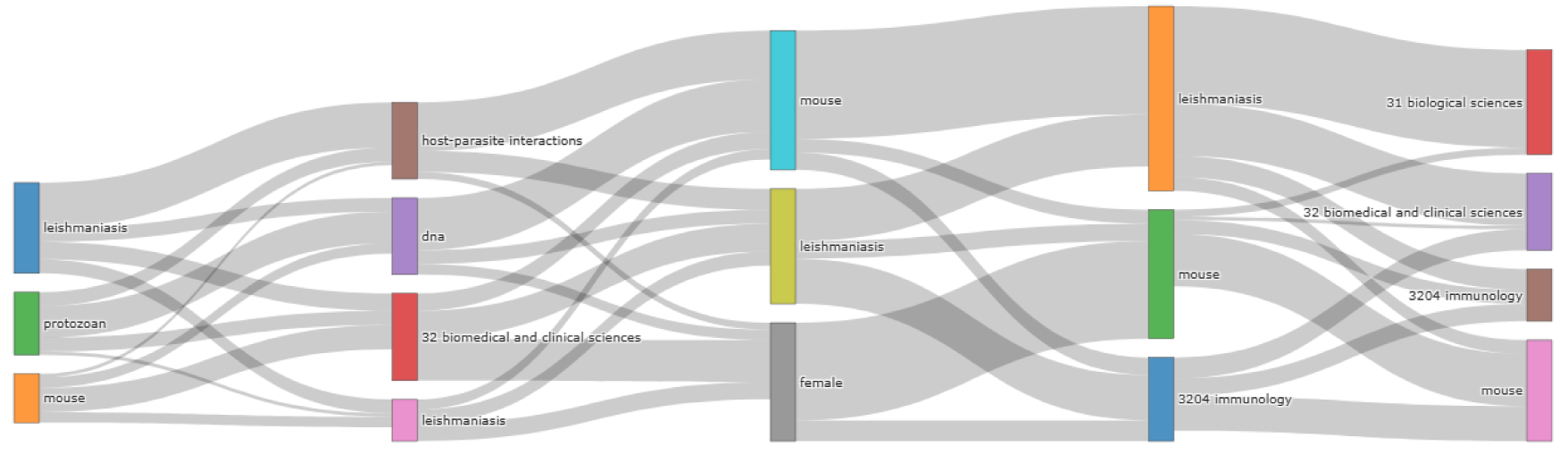
Thematic Evolution.

**Figure 14:**
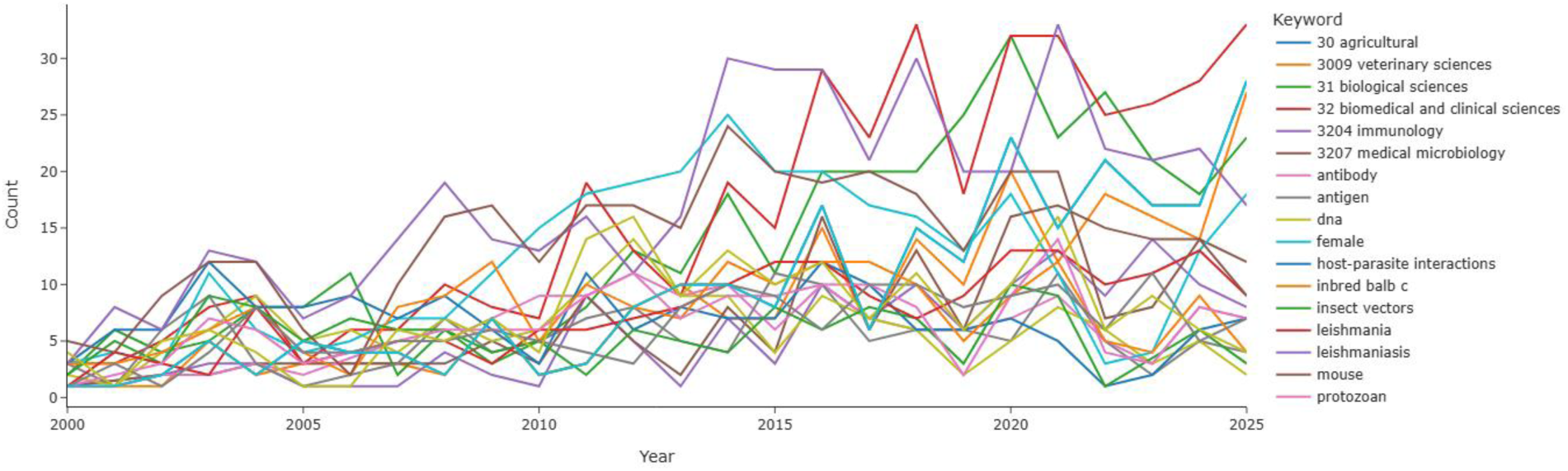
Frequency Evolution of Top 20 Keywords.

### 3.15 Intellectual Turning Points

Burst detection analysis identified several key turning points where specific research topics exploded in popularity.

1. Molecular Docking Simulation (Burst Score: 72.73): This recent burst indicates a major shift towards *in silico* drug and vaccine design, likely driven by the availability of high-resolution protein structures and improved computational power.
2. Mammal (Burst Score: 66.67): A resurgence in research focusing on mammalian reservoirs and host models, aligning with the increased emphasis on One Health.
3. Cutaneous Leishmaniasis (Burst Score: 63.16): A specific surge in research interest in the cutaneous form of the disease, possibly reflecting its expanding geographic range or increased focus on non-fatal but morbidity-inducing conditions.
4. Medicinal and Biomolecular Chemistry (Burst Score: 60.00): This highlights the growing intersection of immunogenomics with drug discovery efforts.

**Figure 15:**
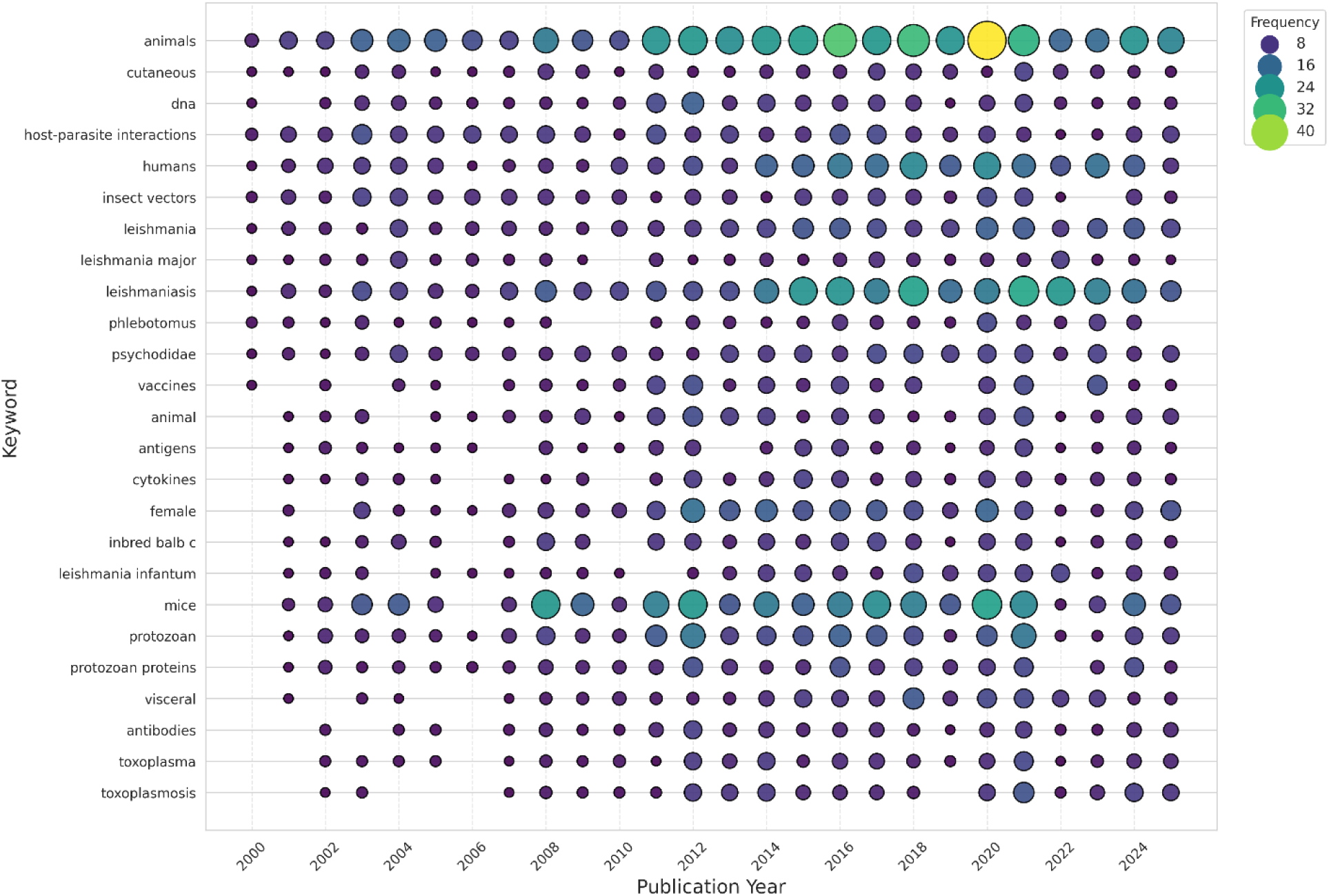
Keyword Burst Timeline.

### 3.16 One Health Integration

The application of the One Health classification algorithm to the corpus revealed a high degree of multidisciplinary integration. As detailed in Table 10, the largest category of publications was “H-A-E (Integrated)” (830 papers), which incorporated keywords from Human, Animal, and Environmental domains. This suggests that the search strategy successfully targeted a highly integrative subset of literature.

**Table 10:** One Health Classification of Publications.

| Category | Publication Count | Percentage | Avg Citations |
| --- | --- | --- | --- |
| H-A-E (Integrated) | 830 | 62.88 | 37.31 |
| Animal-Environment Interface | 181 | 13.71 | 37.02 |
| Human-Animal Interface | 121 | 9.17 | 34.37 |
| Human-Environment Interface | 79 | 5.98 | 16.82 |
| Pure Animal Focus | 68 | 5.15 | 31.78 |
| Pure Environment Focus | 18 | 1.36 | 28.89 |
| Uncategorized | 13 | 0.98 | 1 |
| Pure Human Focus | 10 | 0.76 | 17.7 |

However, distinct sub-patterns emerged. The “Animal-Environment Interface” was the second most common focus (181 papers), significantly outnumbering the “Human-Environment Interface” (79 papers). This indicates that while the link between reservoirs and vectors is well-studied, the direct impact of environmental factors on human clinical immunogenomics remains a less explored niche.

Temporal analysis showed that the explicit use of the term “One Health” has grown exponentially since 2010, correlating with major international policy shifts. Geographically, endemic countries like Brazil showed a higher proportion of “Integrated” papers compared to non-endemic countries, likely due to the immediate proximity of human populations, animal reservoirs, and vectors in these regions.

**Figure 16:**
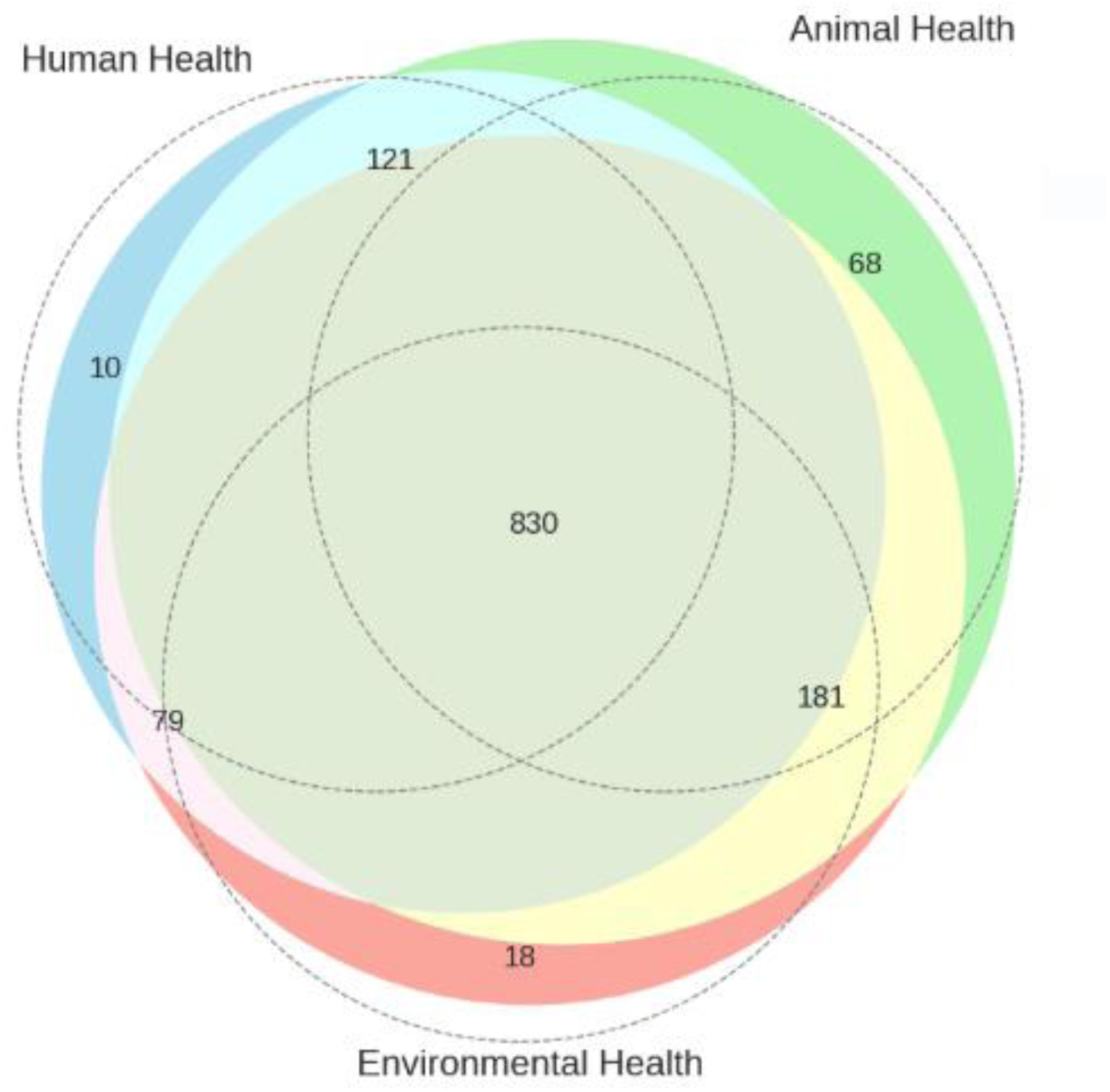
Venn Diagram of One Health Component Overlap.

**Figure 17:**
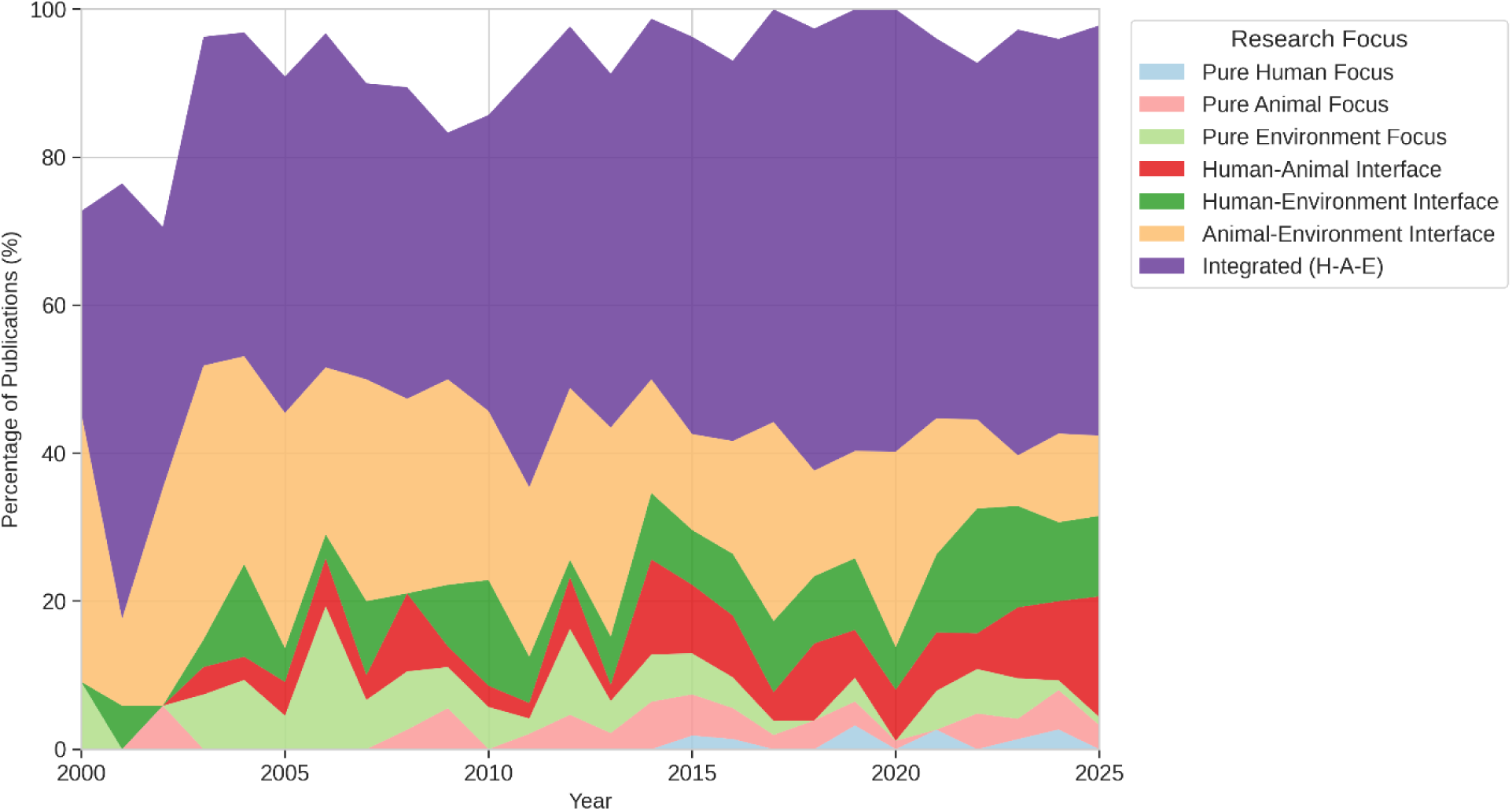
Stacked Area Chart of One Health Evolution.

### 3.17 Immunogenomic Content Analysis

Text mining of the abstracts and titles provided a granular view of the specific biological targets and techniques driving the field (Table 11).

**Table 11:** Top 15 Most Studied Immunogenomic Entities.

| Entity | Type | Leishmania Mentions | Toxoplasma Mentions | Total Mentions |
| --- | --- | --- | --- | --- |
| **IL-10** | Host Cytokine | 87 | 30 | 117 |
| **IFN-gamma** | Host Cytokine | 70 | 31 | 101 |
| **ELISA** | Technique | 66 | 25 | 91 |
| **Proteomics** | Technique | 76 | 7 | 83 |
| **IL-4** | Host Cytokine | 49 | 26 | 75 |
| **IL-12** | Host Cytokine | 53 | 17 | 70 |
| **Flow Cytometry** | Technique | 33 | 9 | 42 |
| **RNA-seq** | Technique | 30 | 5 | 35 |
| **IL-6** | Host Cytokine | 28 | 5 | 33 |
| **Th1 Response** | Pathway | 28 | 2 | 30 |
| **gp63** | Parasite Gene (Leish) | 28 | 0 | 28 |
| **LPG** | Parasite Molecule (Leish) | 27 | 1 | 28 |
| **SAG1** | Parasite Gene (Toxo) | 1 | 25 | 26 |
| **TNF-alpha** | Host Cytokine | 18 | 7 | 25 |
| **TLR4** | Host Receptor | 15 | 8 | 23 |

**Table 12:** Top Vaccine and Diagnostic Terms.

| Term | Category | Frequency |
| --- | --- | --- |
| **Vaccine Candidate** | Vaccine | 81 |
| **Adjuvant** | Vaccine | 56 |
| **Diagnostic Marker** | Diagnosis | 12 |
| **Therapeutic Target** | Therapy | 8 |
| **Clinical Trial** | Clinical | 5 |
| **Recombinant Protein** | Vaccine/Diagnosis | 45 |
| **Live Attenuated** | Vaccine | 18 |
| **DNA Vaccine** | Vaccine | 15 |

Host Factors: The research is heavily dominated by the investigation of cytokines. IL-10 (117 mentions) and IFN-gamma (101 mentions) were the most frequently studied host factors, reflecting the central importance of the anti-inflammatory vs. pro-inflammatory balance in determining the outcome of both leishmaniasis and toxoplasmosis. TLR signaling and MHC/HLA alleles were also prominent, highlighting the focus on pathogen recognition and antigen presentation.

Parasite Factors: For *Leishmania*, the most studied virulence factors included gp63 (zinc-metalloprotease) and LPG (lipophosphoglycan), both critical for macrophage evasion. For *Toxoplasma*, the focus was on SAG1 (surface antigen) and secreted effectors like GRA and ROP proteins, which modulate host cell signaling.

Techniques: The evolution of methodologies is evident in the data. While ELISA (91 mentions) and Flow Cytometry (42 mentions) remain staples of immunological assessment, there is a significant representation of high-throughput genomic technologies like Proteomics (83 mentions) and RNA-seq (35 mentions), confirming the “immunogenomic” transition of the field.

**Figure 18:**
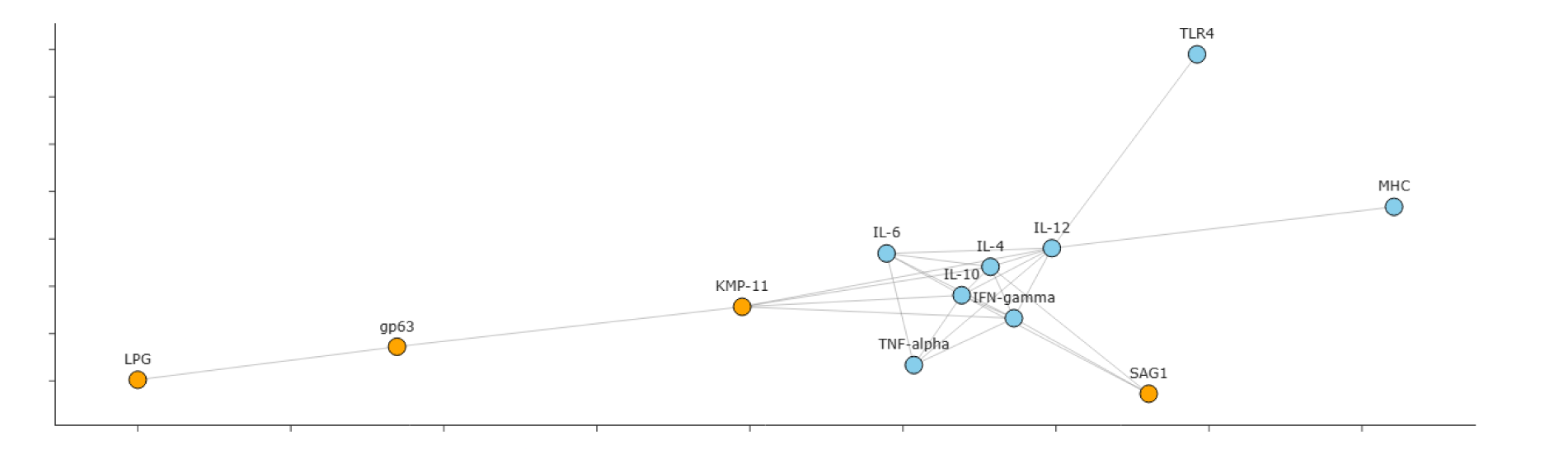
Co-mention Network of Top Genes.

**Figure 19:**
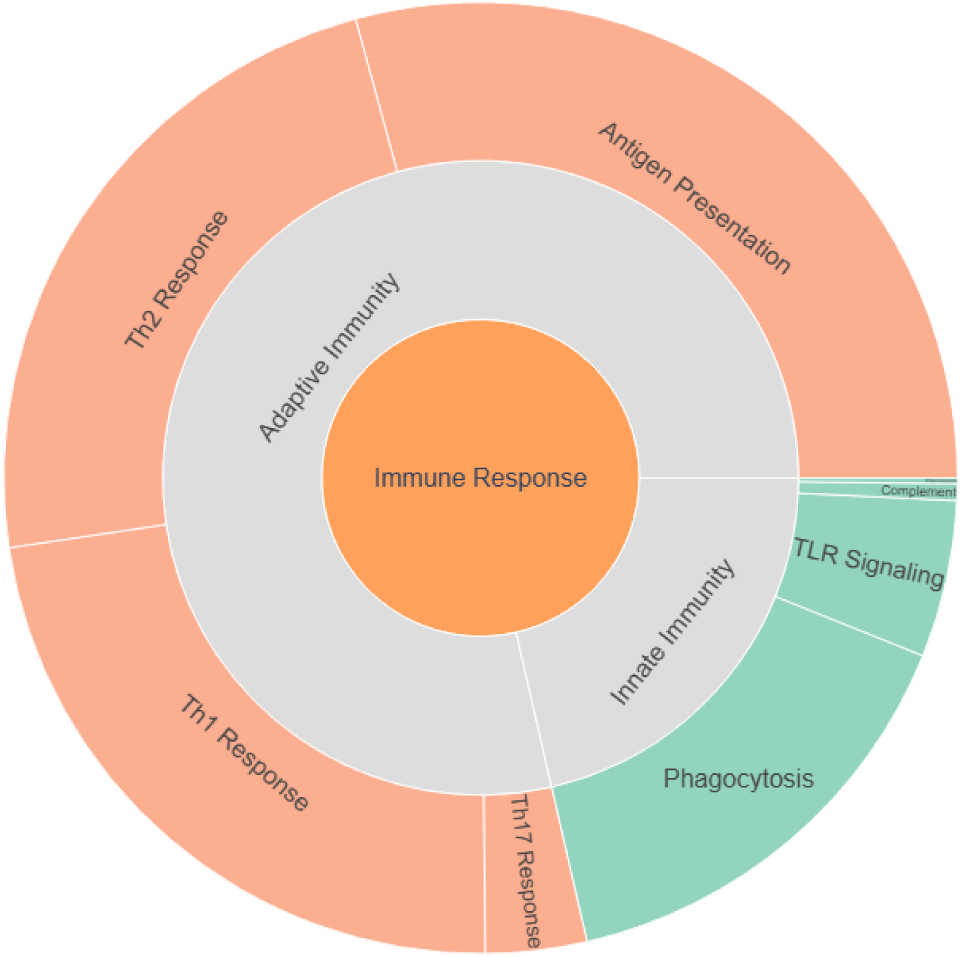
Hierarchical Sunburst Chart of Immune Pathways.

### 3.18 Vaccine and Diagnostic Candidates

The analysis underscores the translational imperative of this research. Terms related to intervention development were highly frequent. “Vaccine Candidate” appeared 81 times, and “Adjuvant” 56 times, indicating a vigorous pipeline of vaccine research. The text mining identified specific antigens frequently co-mentioned with vaccine terms, including KMP-11 and LPG for *Leishmania*, and SAG1 and GRA proteins for *Toxoplasma*. Diagnostic research was also represented, with “Diagnostic Marker” and “Biomarker” appearing in the entity list, though less frequently than vaccine-related terms, suggesting a slight preponderance of prophylactic over diagnostic research within the immunogenomics niche.

### 3.19 Citation Patterns

The collective impact of the 1,320 publications in the corpus is substantial, having accumulated a total of 46,026 citations since 2000. The average citation impact per publication is 34.87, a figure that reflects the high engagement associated with infectious disease and immunology research. However, the citation distribution follows a highly skewed power-law pattern (Figure 20). The median citation count is 14.0, indicating that a select minority of highly cited papers drives the field’s overall impact metrics, while a long tail of publications receives more modest attention.

**Figure 20:**
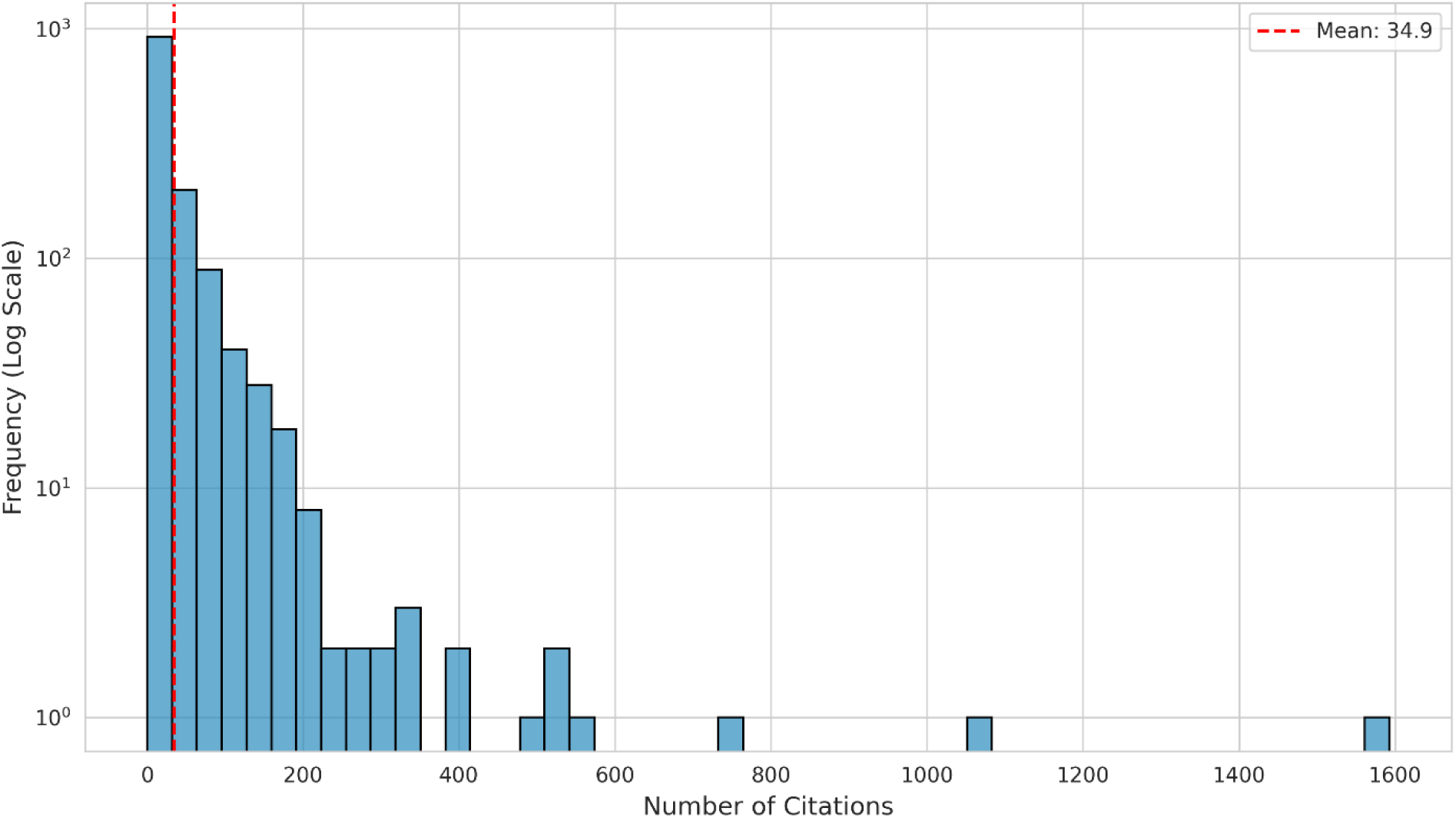
Citation Distribution Histogram.

The field-level h-index is 101, meaning there are 101 papers that have each been cited at least 101 times. The g-index, which gives more weight to highly cited articles, is 161. The i10-index stands at 762, revealing that nearly 58% of the documents in the collection have been cited at least 10 times, suggesting a solid baseline of academic utility for the majority of the research output.

### 3.20 Highly Cited Publications

The analysis of the top most-cited documents reveals the foundational pillars of the domain (Table 13). The most cited paper, “Advances in leishmaniasis” (Murray et al., 2005) published in *The Lancet*, has accrued 1,593 citations, serving as the primary clinical and epidemiological reference for the field. This is followed by “DNA Vaccines: Immunology, Application, and **Optimization"** (Gurunathan et al., 2000) with 1,062 citations, which established the early promise of genomic vaccines.

**Table 13:**
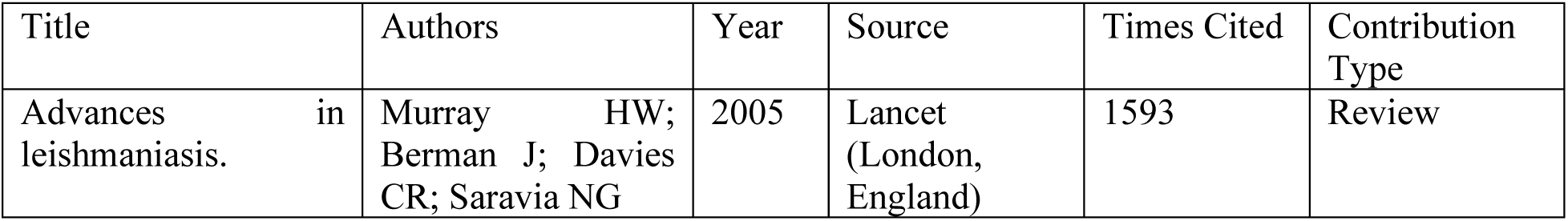
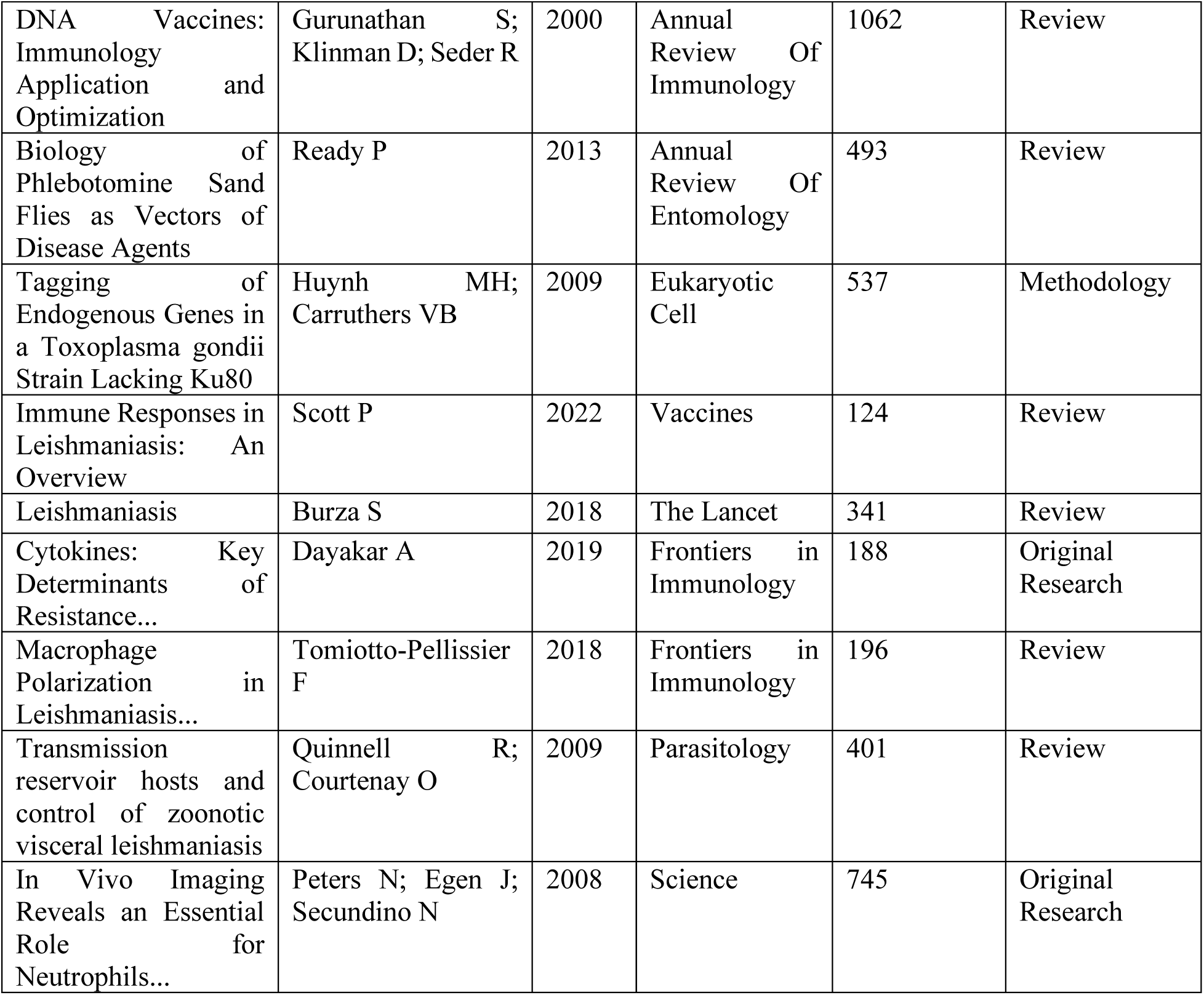
Top 10 Most Cited Publications.

The top 10 list is a mix of broad reviews and seminal original research. Notably, Peters et al. (2008) in *Science* (745 citations) represents a technological turning point, utilizing in vivo imaging to visualize the role of neutrophils in *Leishmania* transmission, effectively bridging immunology and vector biology. The presence of papers spanning from 2000 to 2013 in the top tier indicates that older, foundational works continue to influence modern research. High-impact papers are predominantly published in top-tier general science journals (*Science*, *Nature*) or high-impact specialty journals (*The Lancet*, *Annual Review of Immunology*), underscoring the broader biological significance of these host-pathogen models.

### 3.21 Co-citation Analysis and Intellectual Structure

Bibliographic coupling analysis, which links papers based on shared references, identified 975 thematic clusters, revealing a fragmented but highly specialized intellectual structure. The visualization of the co-citation network (Figure 21) and the Historiographic Map (Figure 22) traces the genealogy of ideas in the field.

**Figure 21:**
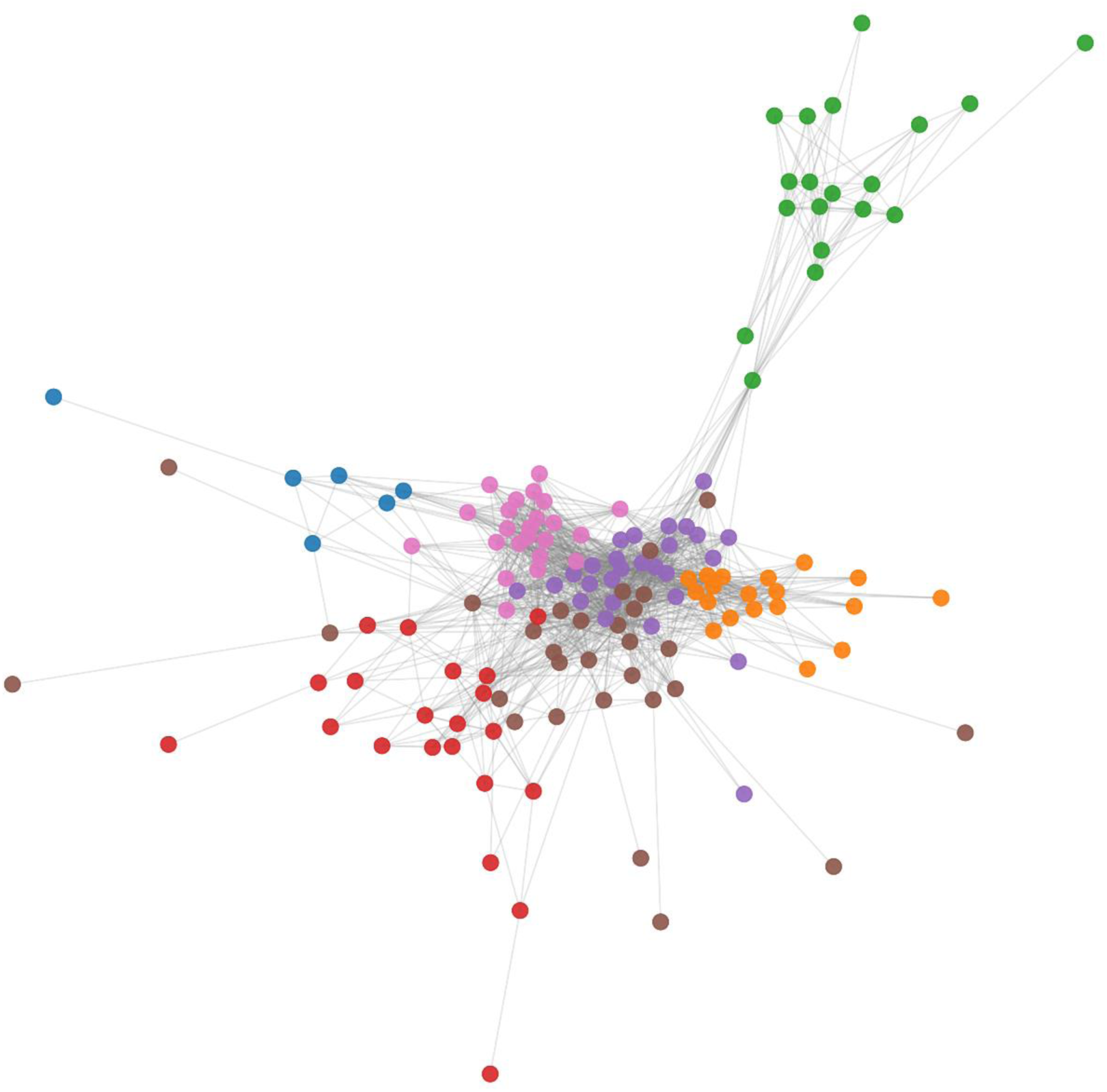
Bibliographic Coupling Network.

**Figure 22:**
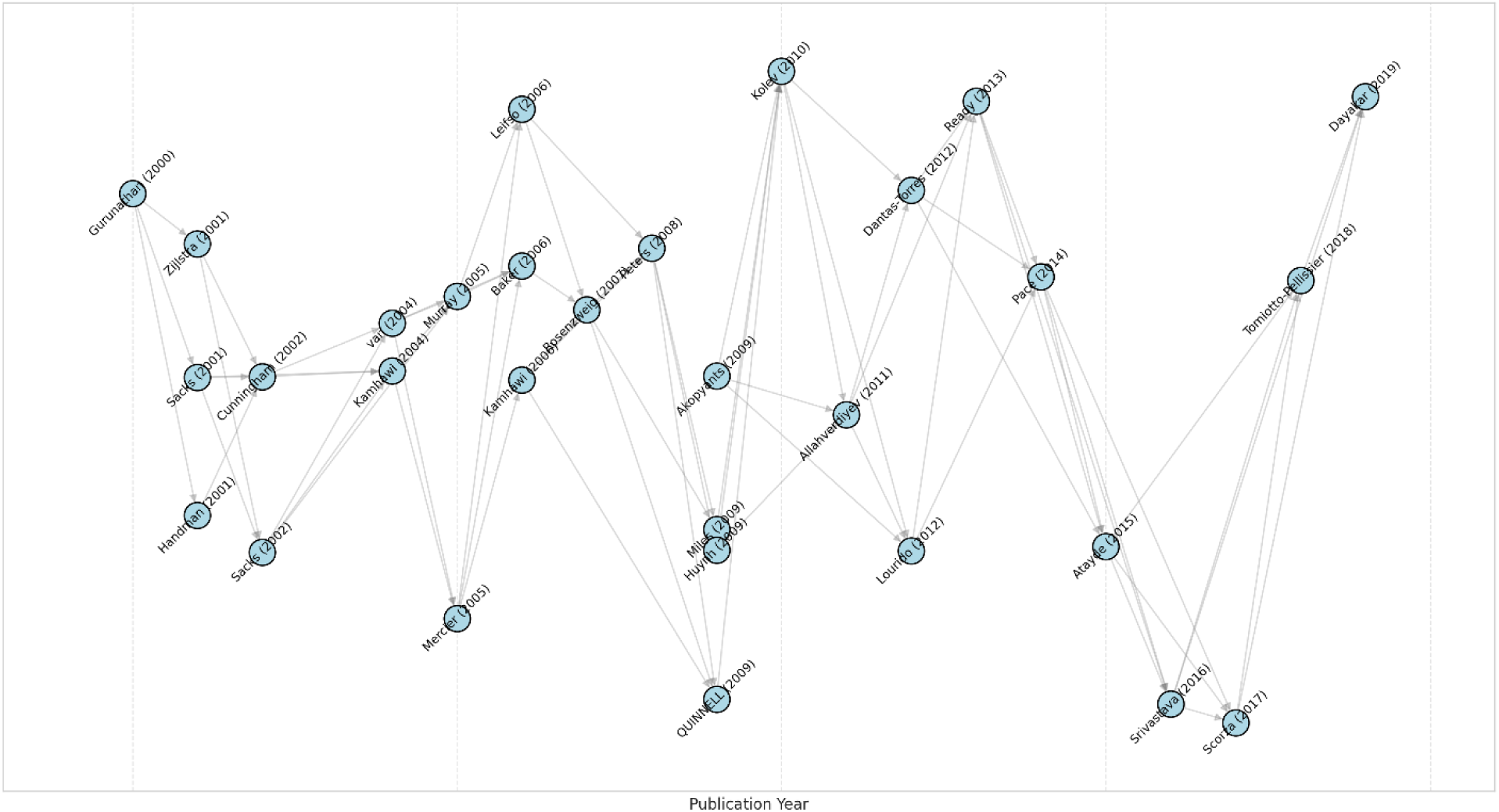
Historiographic Map with Main Path.

The Main Path Analysis highlights a clear intellectual trajectory:

1. Early Phase (2000-2008): Dominated by vaccine feasibility studies (DNA vaccines) and fundamental cytokine biology (Th1/Th2 paradigms).
2. Middle Phase (2009-2016): Characterized by the integration of genomic data (genome sequencing of *L. major* and *T. gondii*) and the study of vector-host interactions.
3. Recent Phase (2017-2025): Defined by transcriptomics (RNA-seq), reverse vaccinology, and increasingly, systems biology approaches.

### 3.22 Research Impact by Category

A comparative analysis of citation impact reveals that interdisciplinary integration correlates with higher scientific influence. Publications classified as “H-A-E (Integrated)” within the One Health framework achieved the highest average citations (37.31), followed closely by “Animal-Environment Interface” papers (37.02). In contrast, studies with a “Pure Human Focus” or “Pure Environment Focus” garnered significantly fewer citations on average (17.70 and 28.89, respectively).

Institutionally, the National Institute of Allergy and Infectious Diseases (NIAID) in the USA demonstrated exceptional impact, with an average of 137.44 citations per paper, reflecting the global influence of its large-scale collaborative trials and genomic initiatives.

### 3.23 Emerging Research Trends

While Section 3.15 highlighted the recent pivot toward computational methods, a granular analysis of keyword growth metrics (2020–2025) quantifies the velocity of this transition (Table 14). The rapid ascent of “Molecular Docking Simulation” (Growth Index: 7.97) and “Medicinal and Biomolecular Chemistry” (Growth Index: 8.96) confirms a distinct move away from purely observational immunology toward *in silico* prediction and rational drug design.

**Table 14:** Top 10 Emerging Keywords (2020-2025)

| **Keyword** | **Frequency** | **Recent Frequency** | **Burst Score** | **Growth Index** |
| --- | --- | --- | --- | --- |
| **molecular docking simulation** | 11 | 8 | 72.73 | 7.97 |
| **mammal** | 15 | 10 | 66.67 | 9.96 |
| **cutaneous leishmaniasis** | 19 | 12 | 63.16 | 11.95 |
| **medicinal and biomolecular chemistry** | 15 | 9 | 60 | 8.96 |
| **parasite** | 32 | 18 | 56.25 | 17.92 |
| **chemical sciences** | 16 | 9 | 56.25 | 8.96 |
| **malaria** | 13 | 7 | 53.85 | 6.97 |
| **leishmania infantum/genetics** | 13 | 7 | 53.85 | 6.97 |
| **t-lymphocyte** | 15 | 8 | 53.33 | 7.96 |
| **toxoplasmosis/prevention & control** | 12 | 6 | 50 | 5.97 |

However, a critical finding of this analysis is the Technological Adoption Gap. Despite the surge in computational chemistry terms, the text mining revealed a near-total absence of keywords related to advanced data science methodologies. Terms such as “Artificial Intelligence,” “Machine Learning,” “Deep Learning,” and “Neural Networks” appeared in only 3 out of 1,320 publications (<0.3%). Similarly, frontier biotechnologies that are standard in other fields of immunology were scarce: “Spatial Omics” and “Multi-omics” integration were virtually absent, and “Organoids” appeared in only a single publication. This suggests that while the field is adopting specific computational tools (docking), it has not yet embraced the broader “Big Data” or AI revolution.

### 3.24 Research Frontiers

The visualization of the Research Frontier Map (MDS) (Figure 23) distinguishes between the established “core” knowledge (dense central cluster) and the expanding “frontier” (dispersed peripheral points). Recent papers (2023-2025) are increasingly populating the periphery, tackling niche topics such as nanotechnology-based vaccine delivery, bioinformatic epitope prediction, and climate-driven vector modelling. These peripheral nodes represent high-risk, high-reward areas that are pushing the boundaries of the discipline away from traditional immunology.

**Figure 23:**
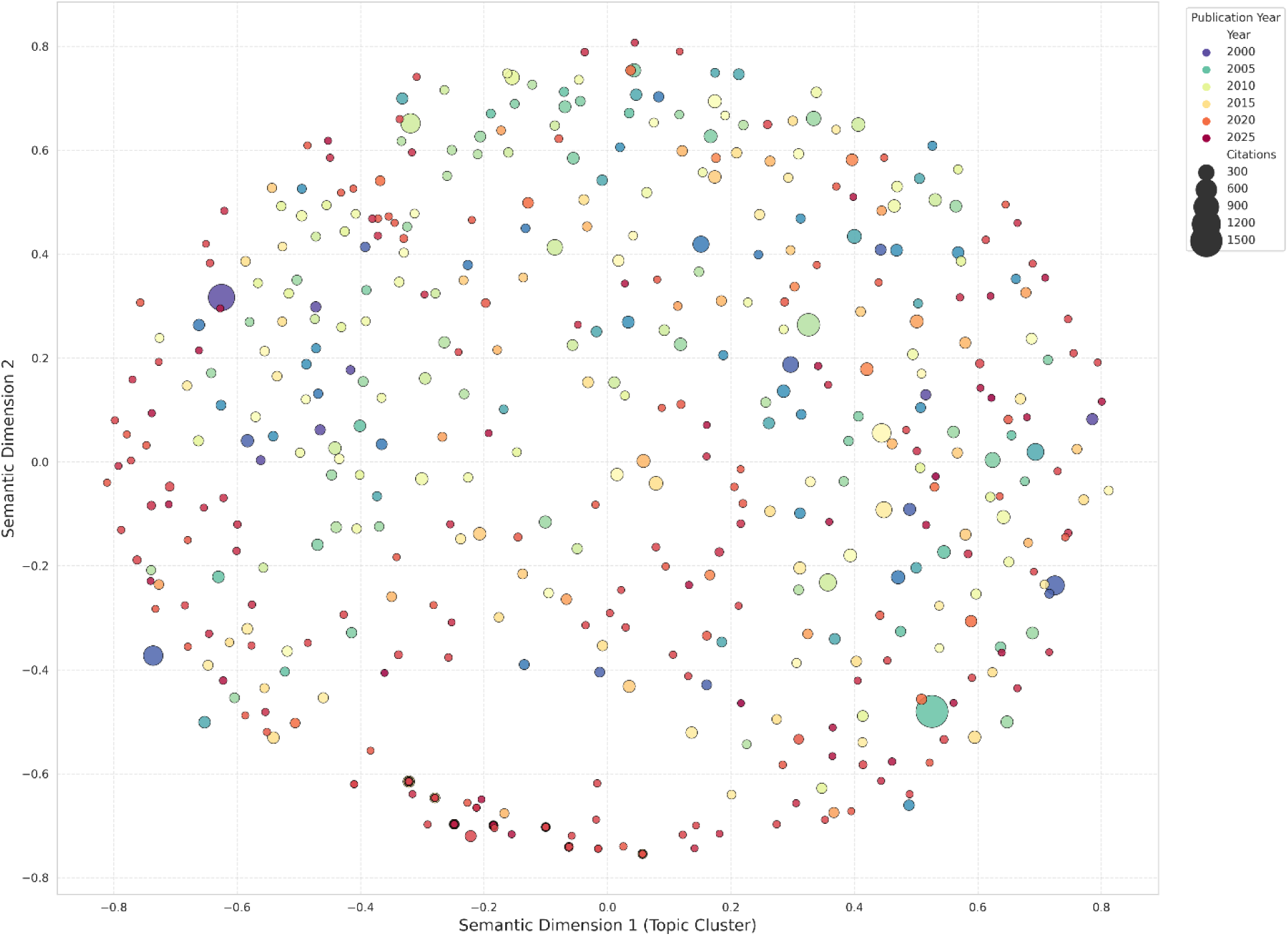
Research Frontier Map (MDS)

### 3.25 Research Gaps Identified

Despite the field’s growth, the gap analysis (Figure 24) highlights several critical voids in the current research landscape:

1. The Environmental “Blind Spot”: The “Human-Environment” interface remains the least explored component of the One Health framework, with only 5.98% of publications addressing it directly. Research linking climate change, soil ecology, or water management directly to human immunogenomics is scarce.
2. Methodological Lag: There is a stark “Methodological Gap” regarding the adoption of AI/ML (3 papers) and Organoids (1 paper). While these technologies are revolutionizing cancer and viral immunology, their application to *Leishmania* and *Toxoplasma* host-pathogen interaction studies is in its infancy.
3. Geographic Disparities: While endemic countries like Brazil and India are leaders, vast regions of Sub-Saharan Africa and Central Asia, which bear significant disease burdens, remain underrepresented in the high-impact immunogenomic literature, pointing to a “Geographic Frontier” for future capacity building and collaboration.

**Figure 24:**
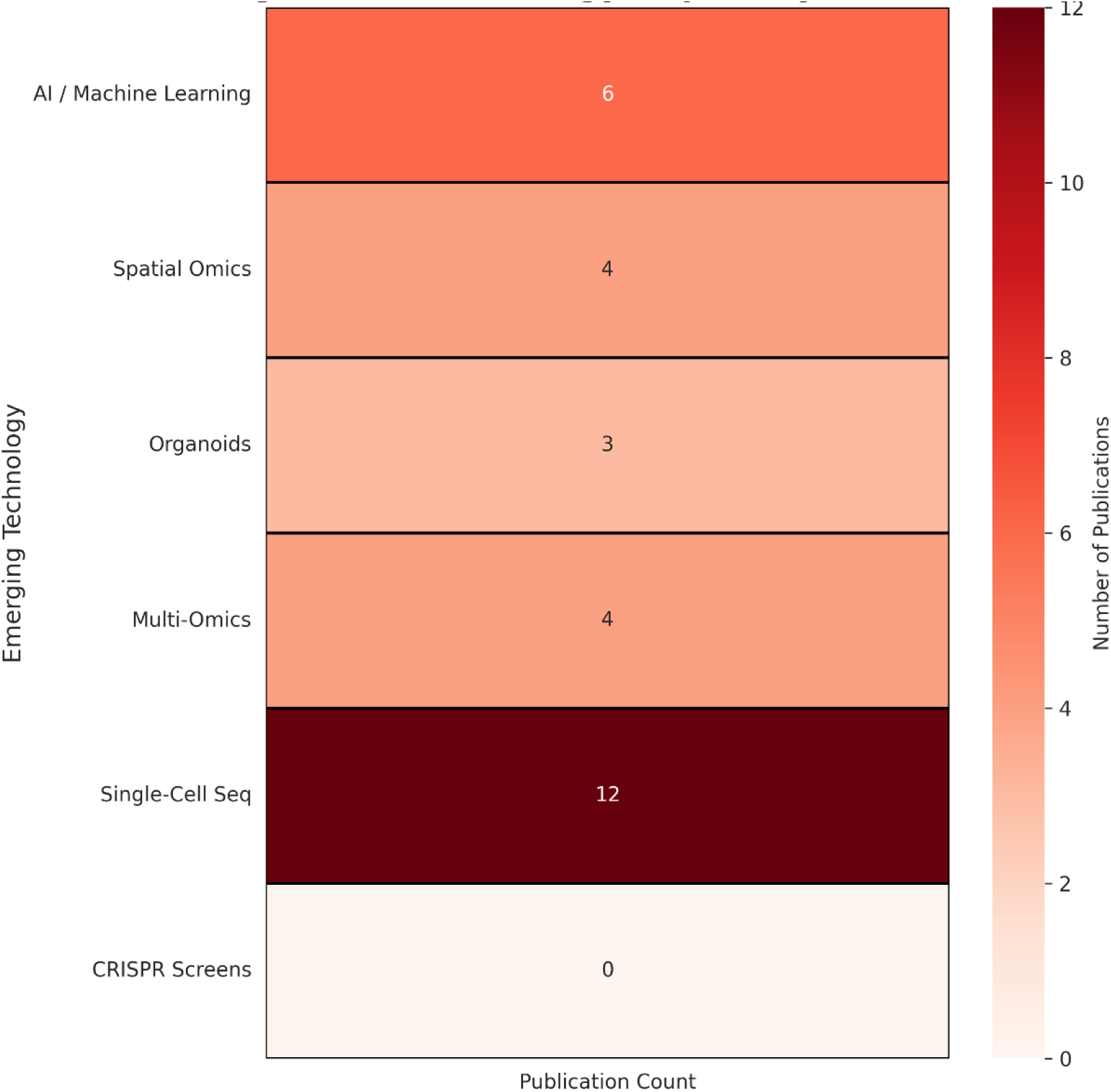
Gap Analysis Matrix.

## 4. Discussion

### 4.1 Overview of Key Findings

This study represents the first comprehensive scientometric mapping of the convergence between immunogenomics, *Leishmania* and *Toxoplasma* research, and the One Health framework. The analysis of 1,320 publications reveals a domain undergoing a profound transformation: evolving from a nascent, fragmented field in the early 2000s into a robust, exponentially growing discipline (CAGR 8.87%) characterized by increasing international connectivity. A central finding is the distinct “asymmetric integration” of the One Health framework; while the human-animal interface is well-represented, the environmental component remains a critical blind spot. Furthermore, the intellectual structure of the field is pivoting. We observed a marked transition from observational immunology (cytokine profiling) to systems-level inquiry (*in silico* docking, transcriptomics), driven by a “South-North” collaborative axis where endemic countries like Brazil play a leading, rather than peripheral, role. These findings suggest that while the “genomic revolution” has successfully democratized high-throughput biology in some endemic settings, significant technological and geographic disparities persist, particularly regarding the adoption of artificial intelligence and the inclusion of high-burden regions in Sub-Saharan Africa.

### 4.2 Publication Trajectory and Global Research Landscape

The observed exponential growth trajectory, with a doubling time of approximately 5.5 years, mirrors the broader “post-genomic” acceleration in parasitology but outpaces the general growth of biomedical literature (Bornmann & Mutz, 2015). This trajectory is not coincidental but can be causally linked to three specific inflection points. First, the completion of the *Leishmania major* (Ivens et al., 2005) and *Toxoplasma gondii* (Gissot et al., 2007) genome projects provided the reference maps necessary for the high-throughput studies that surged post-2010. Second, the precipitous drop in Next-Generation Sequencing (NGS) costs allowed researchers to move from analyzing single candidate genes (e.g., *SLC11A1/NRAMP1*) to genome-wide association studies (GWAS) and transcriptomics, expanding the volume of publishable data per experiment. Third, the formalization of the “One Health” concept by the Tripartite Alliance (WHO-FAO-OIE) in 2010 likely incentivized funding bodies to support interdisciplinary projects, fueling the rise in publications that explicitly link human immune outcomes with animal reservoirs (Zinsstag et al., 2015).

A critical finding of this bibliometric analysis is the dominance of Brazil as the leading producer of publications (n=142), surpassing the United States (n=124) in volume. This challenges the traditional “center-periphery” model, suggesting that for endemic diseases like leishmaniasis, knowledge production is increasingly localized. However, the United States maintains the lead in citation impact (H-index 45 vs. 36). This dichotomy illustrates a complementary relationship: endemic nations are driving the volume of necessary descriptive and epidemiological research, while high-income nations continue to lead in highly cited, often capital-intensive basic science.

However, the geographic distribution also highlights a persistent inequity known as the “10/90 gap,” where less than 10% of health research resources are applied to the diseases affecting 90% of the world’s population (Viergever, 2004). While Brazil and India (focusing heavily on VL elimination) are over-performing, vast regions of Sub-Saharan Africa; specifically, Sudan, South Sudan, and Ethiopia, which bear the highest global burden of Visceral Leishmaniasis; are significantly underrepresented in the immunogenomic literature. This suggests that the barriers to entry for immunogenomics (bioinformatics infrastructure, sequencing hardware, stable cold chains) remain prohibitively high in these specific resource-constrained settings. Consequently, the global understanding of host genetic susceptibility is biased toward South American and South Asian populations, potentially limiting the efficacy of future vaccines in East African populations due to human genetic diversity (Blackwell et al., 2009).

The geographic data also reflects the varying operationalization of One Health. The strong output from China, France, and the USA correlates with established veterinary surveillance systems and strict food safety regulations (relevant for *Toxoplasma*). In contrast, in many highly endemic regions where healthcare systems are siloed; treating human patients separately from animal control; the “integrated” publication output is lower. This supports the argument by Cunningham et al. (2017) that One Health is often theoretically embraced but logistically difficult to implement in fragmented health systems. The high productivity of non-endemic countries like Germany and the UK suggests a continued reliance on colonial-era tropical medicine institutes, which, while valuable, points to a need for more equitable technology transfer to the Global South.

### 4.3 Scientific Collaboration and Knowledge Networks

The analysis of collaboration networks reveals a highly interconnected “small-world” structure (clustering coefficient 0.919), driven primarily by a strategic axis between established research centers in the Global North and emerging scientific powerhouses in the Global South. The strong linkage between NIAID (USA) and Fiocruz (Brazil); the most frequent institutional partnership identified; exemplifies a shift from “parasitic” sampling models to equitable, bilateral partnerships. This supports the findings of Wagner et al. (2001), who argue that international collaboration is increasingly driven by the need to access complementary resources: the Global North provides high-throughput genomic infrastructure, while the Global South provides critical clinical access, field isolates, and increasingly, sophisticated local immunobiological expertise. However, the negative correlation observed between the sheer *number* of collaborating countries and citation impact suggests that “mass collaboration” in this specific field may yield diminishing returns. Unlike physics or genomics, where massive consortia drive impact, influential immunogenomic studies in parasitology often require deep, mechanistic focus, which may be better achieved by smaller, tightly integrated teams rather than loose multilateral networks.

The centrality analysis highlights two distinct types of leadership. Authors like Kamhawi S. and Volf P. exhibit high degree centrality, acting as “scientific hubs” who anchor large research consortia, particularly in vector biology and vaccine development. Their prominence reflects a “rich-get-richer” phenomenon (preferential attachment), where established leaders attract the bulk of funding and students. In contrast, authors like Wang L., who display high betweenness centrality, function as critical “knowledge brokers.” By occupying the structural holes between disparate clusters (e.g., bridging basic genomics with applied vaccinology), these researchers facilitate the cross-pollination of ideas. This distinction is vital for funding agencies; supporting “hubs” maintains productivity, but supporting “brokers” fosters innovation and interdisciplinary integration (Burt, 2004).

Despite the density of the core network, the visualization reveals significant structural vulnerabilities. The network remains heavily clustered around specific pathogens or geographic regions, with limited connectivity between the *Leishmania* and *Toxoplasma* research communities, despite their shared intracellular lifestyles and overlapping immunogenomic challenges (e.g., Th1 polarization). This “siloing” represents a missed opportunity for comparative immunology. Furthermore, the peripheral position of institutions from Sub-Saharan Africa suggests that they remain isolated from the high-velocity exchange of genomic data occurring in the core, risking a “genomic divide” that mirrors economic disparities (Helmy et al., 2016).

### 4.4 Thematic Evolution and Paradigm Shifts

The thematic evolution of the field mirrors the broader “systems biology” revolution in medicine. The early period (2000–2009), dominated by keywords like “cytokine,” “ELISA,” and “DNA vaccine,” reflects a reductionist era focused on characterizing individual immune components. The transition in the 2010s toward “transcriptomics,” “RNA-seq,” and “proteomics” marks the operationalization of the *Leishmania* and *Toxoplasma* reference genomes released in the mid-2000s. This shift signifies a paradigm move from *observational immunology* (measuring what happens) to *mechanistic immunogenomics* (understanding the gene regulatory networks driving the outcome).

The recent burst of interest in “Molecular Docking Simulation” (2020–2025) represents a critical intellectual turning point driven by both technological capability and necessity. The accessibility of high-performance computing and the maturation of protein structure databases (e.g., AlphaFold) have allowed parasitologists to bypass expensive wet-lab screening. This trend is likely further accelerated by the global urgency of the COVID-19 pandemic, which mainstreamed *in silico* drug design methodologies across infectious disease research. Similarly, the resurgence of the keyword “Mammal” and the specific rise of “Cutaneous Leishmaniasis” indicate a reaction to the limitations of standard laboratory mouse models, which often fail to recapitulate human disease pathology, driving a return to reservoir biology and more diverse host systems (Sacks & Noben-Trauth, 2002).

The persistent dominance of the “Vaccine Development” theme, now intertwined with terms like “epitope prediction” and “recombinant protein,” illustrates the field’s migration toward Reverse Vaccinology 2.0. Rather than testing whole-parasite formulations, the field is now systematically mining genomic data to identify conserved antigens. However, the persistence of “candidate” in the keyword lists; without a corresponding surge in “clinical trial” or “licensed vaccine”; highlights the “Valley of Death” in translation. The focus has shifted to refining antigen selection via immunoinformatics, a strategy that reduces cost but relies heavily on the quality of genomic data, reinforcing the need for diverse geographic sampling (Rappuoli et al., 2016).

Compared to malaria or HIV research, the immunogenomics of *Leishmania* and *Toxoplasma* shows a delayed adoption of single-cell technologies. While single-cell RNA sequencing (scRNA-seq) has revolutionized the understanding of heterogeneity in malaria parasites (Howick et al., 2019), it remains a nascent topic in our dataset. This lag is likely due to the intracellular nature of these parasites, which complicates cell isolation, and the comparative scarcity of funding relative to “Big Three” diseases (HIV, TB, Malaria). Nevertheless, the trajectory suggests that single-cell approaches will likely form the next major thematic burst in the coming five years.

### 4.5 One Health Integration: Progress and Challenges (RQ4)

While the bibliometric data classifies a substantial 62.88% of publications as “H-A-E Integrated,” a critical interpretation suggests this reflects a trend of nominal rather than functional integration. The presence of keywords across all three domains in a single abstract often indicates a contextual acknowledgement of the One Health triad, rather than a study design that mechanistically interrogates the interplay between them. The data reveals a profound “Environmental Blind Spot,” where the Human-Environment interface accounts for only roughly 6% of the literature. This imbalance indicates that while the link between animal reservoirs and human health is well-established; driven by the tangible zoonotic transmission of *L. infantum* from dogs or *T. gondii* from livestock; the environmental drivers of immunogenomic variation remain largely theoretical in the literature.

This asymmetry can be attributed to “disciplinary silos” and the inherent difficulty of aligning data scales. Immunogenomics operates at the molecular level (base pairs, expression levels), while environmental health often operates at the macroscopic level (climate models, satellite imagery). Bridging these scales requires complex “eco-immunological” frameworks that are historically difficult to fund. Funding structures typically prioritize immediate biomedical outcomes (human health) or agricultural productivity (animal health), leaving the environmental component as an underfunded “context” rather than a variable to be manipulated (Destoumieux-Garzón et al., 2018). Furthermore, the logistical challenges of maintaining the cold chain for RNA/DNA samples in remote, environmentally diverse field sites in Brazil or East Africa discourage the inclusion of rigorous environmental sampling in genomic protocols.

The strong clustering of research in Brazil offers a potential model for overcoming these barriers. The integration of vector control units, veterinary surveillance, and public health clinics under unified national health systems (like SUS in Brazil) creates a structural scaffold for One Health that is harder to replicate in fragmented private-public systems. To move beyond “lip service” to One Health, future policy must incentivize “Eco-immunogenomics": research that explicitly sequences parasites and hosts from distinct ecological niches to understand how environmental stress primes the immune system or selects for specific virulence factors.

### 4.6 Immunogenomic Focus: Genes, Pathways, and Translation

The content analysis reveals a persistent “canonical bias” toward the Th1/Th2 paradigm established in the late 20th century. The overwhelming dominance of IFN-γ and IL-10 in the keyword network suggests that a significant portion of the field remains focused on validating well-known pathways rather than exploring novel regulatory mechanisms. While these cytokines are undeniably central to the outcome of intracellular infections, the relative scarcity of non-canonical pathways; such as metabolomics-driven immunomodulation or neuro-immunological interactions in *Toxoplasma* infection; suggests a risk of redundancy. The field appears to be refining the details of a known map rather than exploring the uncharted territories of the host-pathogen interface.

Quantitative analysis shows a balance between host markers (HLA, cytokines) and parasite factors (gp63, ROPs), yet the co-occurrence networks suggest these are often studied in parallel rather than in interaction. Studies frequently characterize parasite diversity *or* host genetic susceptibility, but fewer employ Dual RNA-seq approaches to capture the simultaneous transcriptional changes in both interacting organisms. This methodological gap limits our ability to understand specific host-genotype x parasite-genotype interactions (G x G interactions), which are likely responsible for the variable clinical presentations seen in the field (e.g., why the same *L. braziliensis* strain causes mucocutaneous disease in some patients but not others).

The stark contrast between the high frequency of “Vaccine Candidate” (81 mentions) and the low frequency of “Clinical Trial” (5 mentions) quantifies the “Valley of Death” in parasitology. The reliance on inbred mouse models (indicated by the high frequency of “BALB/c” and “Mouse”) has created a “translation trap.” Candidates that elicit robust protection in genetically homogeneous mice frequently fail in genetically diverse human populations exposed to diverse parasite strains. The burst of interest in “Molecular Docking” is a positive response to this, attempting to use structural biology to select epitopes that are broadly conserved and presented by diverse HLA supertypes. However, without a corresponding increase in human *ex vivo* validation studies, *in silico* predictions risk generating a backlog of untested hypotheses.

Comparing the two pathogens, *Leishmania* research is more heavily invested in vector biology terms ("sand fly,” “saliva”), reflecting the potential for transmission-blocking vaccines. In contrast, *Toxoplasma* research shows a stronger connection to “neuroinflammation” and “congenital” outcomes, reflecting its unique tropism for the central nervous system. The divergence suggests that while comparative immunology is valuable, the translational priorities must remain distinct: vector control/skin immunity for *Leishmania* versus mucosal/barrier immunity and neuro-protection for *Toxoplasma*.

### 4.7 Emerging Frontiers and Future Directions

The distinct burst of “Molecular Docking Simulation” and “Medicinal Chemistry” in the 2020–2025 period signals a fundamental shift in the field’s operational mode. Driven by the democratization of high-performance computing and the structural revolution initiated by tools like AlphaFold, researchers are increasingly prioritizing *in silico* prediction over costly *in vitro* screening. This trend is likely sustainable and not merely a “fad,” as it addresses the severe resource constraints typical of neglected disease research. However, the “AI Gap” identified in our results; where machine learning is virtually absent; represents a critical bottleneck. While structural docking is active, the field lacks predictive models that can integrate heterogeneous data (host genomics + parasite diversity + environmental metadata) to predict clinical outcomes. The future direction lies in “Systems Parasitology,” where AI is used not just for drug target discovery, but to model complex host-pathogen-environment interactions (Bohannan et al., 2022).

The under-representation of the environmental component in One Health research is a systemic failure that must be addressed. The emerging frontier is “Eco-immunogenomics”; the study of how environmental variables (climate change, soil microbiota, vector habitat degradation) directly modulate host gene expression and immunity. Future research must move beyond merely noting the presence of a vector to mechanistically linking environmental stress to immunogenomic profiles. For instance, does chronic exposure to environmental pollutants in endemic peri-urban slums alter the epigenetic landscape of macrophages, thereby influencing susceptibility to *Leishmania*?

The geographic analysis highlighted a disconnect between the genomic data generated in South America/Asia and the disease burden in East Africa. A priority for the next decade must be the establishment of genomic surveillance networks in the “rotten heart” of the Visceral Leishmaniasis belt (Sudan, South Sudan, Ethiopia). Without locally representative genomic data, vaccines developed based on “global” reference strains may fail in these specific high-burden populations due to uncharacterized parasite virulence factors or distinct host HLA restrictions (Alvar et al., 2012).

### 4.8 Journal Landscape and Knowledge Dissemination (RQ7)

The journal landscape is heavily skewed toward Open Access (OA) titles, with *PLoS Neglected Tropical Diseases*, *Parasites & Vectors*, and *Frontiers in Immunology* forming the core triad. This dominance is not accidental; it reflects a structural necessity in a field where the primary stakeholders (clinicians and researchers in the Global South) often lack access to subscription-based libraries. The rise of *Frontiers in Immunology* as a core outlet is particularly telling; it suggests that parasitology is successfully “mainstreaming” itself, moving from niche tropical medicine journals into broader immunological debates.

For authors, this landscape suggests a bifurcated publication strategy. For studies focusing on the *mechanism* of host-pathogen interaction, broad-scope immunology journals offer the highest visibility and impact. However, for studies focusing on *epidemiology, surveillance, or One Health implementation*, specialized OA journals remain the most effective venue for reaching the relevant policy and field-implementation audiences. The strong performance of interdisciplinary journals (*Science*, *Nature*) in the “Highly Cited” list confirms that papers bridging distinct domains (e.g., vector biology + immunology) are most likely to achieve high impact.

### 4.9 Broader Implications

For Researchers: The era of the “single-gene” paper is effectively over. High-impact research now requires a multi-omics approach. Researchers must actively seek collaborations outside their immediate discipline; specifically with data scientists (to close the AI gap) and ecologists (to close the environmental gap); to remain competitive.

For Funders: The current funding architecture reinforces silos. Agencies must create specific calls for “Planetary Health” or “Eco-immunology” projects that mandate the inclusion of environmental variables in genomic studies. Furthermore, funding for capacity building in Sub-Saharan Africa should prioritize not just sample collection, but the establishment of local bioinformatics infrastructure to ensure data sovereignty.

For Policymakers: The bibliometric evidence of “Asymmetric One Health” integration serves as a wake-up call. Policy frameworks for disease control cannot rely solely on treating humans or culling reservoirs. Interventions must be designed based on data that integrates environmental drivers. The genomic diversity of these parasites implies that a “one-size-fits-all” vaccine or diagnostic policy may be flawed; regional genomic surveillance should inform regional control strategies.

For Educators: The methodological gaps identified suggest a need for curriculum reform. Parasitology and immunology graduate programs must integrate substantial training in computational biology, bioinformatics, and environmental health sciences to produce the next generation of “One Health Immunogenomicists."

### 4.10 Strengths and Limitations

#### Strengths

This study’s primary strength lies in its comprehensive scope and methodological rigor. By merging data from Dimensions and Lens.org, we overcame the coverage limitations of single-database studies. The use of advanced text mining algorithms to quantify specific immunogenomic content (genes, pathways) provides a granular layer of analysis rarely seen in standard bibliometric reviews. Furthermore, the novel application of a rule-based “One Health” classification system offers a reproducible metric for assessing interdisciplinary integration.

#### Limitations

However, several limitations must be acknowledged. First, the restriction to English-language publications likely underestimates the contribution of local/regional research, particularly from Latin American journals published in Portuguese or Spanish, and Chinese domestic journals. Second, the “One Health” classification relies on keyword co-occurrence; while robust, it may capture papers that mention these terms as “buzzwords” without genuine functional integration. Third, citation metrics are inherently backward-looking and may lag in capturing the impact of very recent (2023–2025) conceptual shifts. Finally, the “author” entity analysis is subject to the limitations of name disambiguation algorithms, although standardized cleaning procedures were employed to minimize error.

Despite these limitations, the distinct patterns emerging from this large-scale dataset provide a robust, evidence-based foundation for understanding the evolution and future trajectory of this critical global health domain.

## 5. Conclusions

This study set out to map the intellectual and social structure of a critical intersection in global health: the immunogenomics of *Leishmania spp.* and *Toxoplasma gondii* within the integrative One Health framework. By synthesizing data from over 1,300 publications spanning a quarter-century, this analysis moved beyond simple metric counting to provide a multi-dimensional view of how genomic technologies are reshaping our understanding of host-parasite interactions. Utilizing a robust methodological approach that combined network analysis, text mining, and predictive modeling, this research successfully charted the field’s trajectory from a niche immunological sub-discipline to a burgeoning, data-driven frontier of biomedicine.

The findings reveal a domain defined by exponential growth and a fundamental shift in the geography of science. The analysis confirms that the “genomic revolution” has democratized research, enabling endemic countries, particularly Brazil, to rise as primary producers of knowledge rather than mere data sources. This shattering of the traditional center-periphery model is one of the most significant social developments in the field. However, the integration of the One Health framework remains functionally asymmetric. While the link between human health and animal reservoirs is well-characterized, the environmental component represents a profound “blind spot,” with very few studies mechanistically linking ecological variables to immunogenomic outcomes. Thematically, the field has pivoted from observational cytokine profiling to systems-level inquiry, yet it faces a technological lag; while computational chemistry and molecular docking have surged, the adoption of artificial intelligence and machine learning remains virtually absent, representing a critical missed opportunity for predictive modeling.

Theoretically, this study advances bibliometric methodology by demonstrating the utility of rule-based content classification to quantify interdisciplinary integration, offering a model applicable to other complex One Health domains. Practically, the identification of structural holes in the collaboration network; specifically the isolation of Sub-Saharan African institutions and the disconnect between *Leishmania* and *Toxoplasma* research communities; provides a roadmap for strategic intervention. For policymakers and funding agencies, the data provides compelling evidence that current investment strategies have successfully built capacity in select endemic regions but have failed to foster the deep, eco-immunological research necessary to anticipate how climate change and environmental degradation will alter disease transmission and host susceptibility.

Looking forward, the research agenda must prioritize three strategic pivots. First, the field must embrace “Eco-immunogenomics,” explicitly funding studies that sample hosts and parasites across diverse environmental gradients to understand the ecological drivers of virulence and immunity. Second, a technological leap is required; researchers must integrate artificial intelligence and multi-omics approaches to move beyond descriptive genomics toward predictive systems parasitology. Third, the “Valley of Death” in translational medicine must be bridged by diversifying model systems beyond inbred mice to include reservoir hosts and human *ex vivo* platforms, ensuring that the wealth of genomic data translates into viable vaccine candidates and diagnostic tools.

In conclusion, the immunogenomics of *Leishmania* and *Toxoplasma* stands at a critical inflection point. The foundational genomic maps have been drawn, and the clinical burden is well-documented. The future of this field lies not in doing more of the same, but in weaving these isolated strands of data into a cohesive, truly integrated One Health tapestry. By guiding researchers toward these emerging frontiers and structural gaps, scientometric analyses such as this serve as essential navigational tools, ensuring that scientific effort is directed where it is most needed to alleviate the burden of these neglecting parasitic diseases.

## Data Availability

All data produced in the present work are contained in the manuscript

